# Negative Social Ties as Emerging Risk Factors for Accelerated Aging, Inflammation, and Multimorbidity

**DOI:** 10.1101/2025.05.23.25328261

**Authors:** Byungkyu Lee, Gabriele Ciciurkaite, Siyun Peng, Colter Mitchell, Brea L. Perry

**Author notes:** Corresponding author: Byungkyu Lee.

## Abstract

Negative social ties, or “hasslers,” are pervasive yet understudied components of social networks that may accelerate biological aging and morbidity. Using ego-centric network data and DNA methylation-based biological aging clocks (i.e., DunedinPACE and age-accelerated GrimAge2) from saliva from a state representative probability sample in Indiana, we examine how negative social ties are associated with accelerated biological aging and a broad range of health outcomes, including inflammation and multimorbidity. Negative relationships are not rare within close relationships, as nearly 30% of individuals report having at least one hassler in their network. These hasslers tend to occupy peripheral network positions and are more likely to be connected through weak, uniplex ties. Importantly, exposure to negative social ties follows patterns of social and health vulnerability, with women, daily smokers, people in poorer health, and those with adverse childhood experiences more likely to report having hasslers in their networks. Having more hasslers is associated with accelerated biological aging in both rate and cumulative burden: each additional hassler corresponds to approximately 1.5% faster pace of aging and roughly nine months older biological age. Moreover, not all hasslers exert the same influence; kin and non-kin hasslers show detrimental associations, whereas spouse hasslers do not. Finally, a greater number of hasslers is associated with multiple adverse health outcomes beyond epigenetic aging. These findings together highlight the critical role of negative social ties in biological aging as chronic stressors and the need for interventions that reduce harmful social exposures to promote healthier aging trajectories.

## Introduction

Aging is a universal biological process involving the gradual accumulation of unrepaired molecular damage, leading to progressive declines in physiological functions and increased vulnerability to disease (*1*, *2*). Despite its universality, individuals vary substantially in their rates of biological aging, distinct from chronological aging, due to genetic predispositions, as well as environmental and social factors (*3*, *4*). Advancements in epigenetic biomarkers, particularly DNA methylation (DNAm)-based epigenetic clocks, have now made it possible to quantify and study biological aging. Early epigenetic clocks primarily measure DNAm patterns correlated with chronological age; therefore, they are less suitable for health research, whereas newer, second- and third-generation clocks, such as DunedinPACE and GrimAge, reflect aging phenotypes, health risks, and mortality outcomes (*5*, *6*). Recent research demonstrates that accelerated biological aging, as indicated by these advanced DNAm clocks, robustly predicts critical health outcomes, including chronic conditions and mortality risk (*7*). With these critical tools, a crucial next step is to explore the factors that may accelerate or slow biological aging by linking molecular mechanisms with the broader social forces that shape the aging process.

Chronic stress is a well-established driver of biological aging, influencing epigenetic modifications that regulate gene expression and inflammation (*8*). While often conceptualized as sources of support that promote health and well-being (*9*–*12*), social relationships may also function as chronic stressors, contributing to increased allostatic load through repeated stress activation (*13*, *14*). This dual nature of social ties—both protective and harmful—raises important questions about their role in biological aging and the pathogenesis of common morbidities. However, existing research relies on methods that capture only the positive dimensions of social relationships (e.g., social support), often overlooking negative or strained ties (*15*). As a result, there is comparatively little empirical insight into how the dark side of networks contributes to biological aging.

A growing body of research suggests that negative ties—relationships characterized by hostility, strain, or excessive burden, making one’s life difficult—may be prevalent and have lasting health consequences (*16*–*18*). These negative ties, which we call “hasslers,” may accelerate biological aging by mimicking the harmful effects of traditional chronic stressors, such as financial strain or workplace stress (*19*, *20*), and contribute to increased inflammation, compromised immune function, and elevated risk for cardiovascular and other diseases (*21*). Several studies have demonstrated that conflict and hostility in the context of marital relationships are associated with accelerated epigenetic aging (*22*–*24*). However, it is unclear whether such stressful relationships outside of marriage have similar biological effects (*10*, *25*, *26*).

Simultaneously, it is crucial to recognize that what respondents describe as “hassling” does not uniformly represent relational toxicity (*19*, *20*). As classic social theory reminds us, conflict and closeness often coexist within intimate ties. This dualistic argument has been articulated by Simmel (*27*): the greatest tensions frequently emerge within the most intimate relationships, precisely because their emotional security allows for friction without dissolution. In this light, some forms of hassling may reflect informal social control born out of love and concern, particularly within spousal or caregiving contexts. Yet, other forms of hassling may operate as chronic stressors, producing role strain and emotional exhaustion, especially within kin networks where obligations are enduring and less easily renegotiated. In contrast, hassling from non-kin ties may be experienced as less consequential or easier to disengage from. Building on theories of social balance and role systems (*27*–*29*), this study examines how the meaning and consequences of “hassling” differ across relationship types with a focus on the implications of such interactions depending on their relational and structural contexts.

These gaps in the literature are due in large part to methodological limitations, such as insufficient data on biological indicators of aging and social networks. To address these gaps, our study draws on an age-heterogeneous, state-representative probability sample in Indiana (i.e., the Person-to-Person Study) that uniquely includes both biomarkers and extensive measures of positive and negative interactions within personal networks using an in-depth ego-centric network module. To assess biological aging, we estimate second- and third-generation epigenetic clocks (age-accelerated GrimAge2 and DunedinPACE) derived from high-quality saliva samples. We begin by documenting the prevalence of negative ties (or *hasslers*) and identifying which individuals are most likely to report them. We then evaluate how having such negative ties is associated with accelerated biological aging, with particular attention to heterogeneity across relationship types (i.e., partners, kins, non-kins). To explore mechanisms, we further investigate the relational characteristics that define these negative ties. In addition, we assess whether negative ties are linked to a wider range of health indicators beyond epigenetic aging and, finally, conduct a series of sensitivity analyses to evaluate the robustness of our findings.

Our main hypothesis is that the presence of hasslers in core (i.e., close) networks is associated with faster biological aging and poorer health; however, such associations do not, on their own, establish a causal effect of negative social ties on aging processes. Several alternative mechanisms could also generate the observed patterns. One is reverse causation: individuals experiencing accelerated biological aging may become more irritable, thereby eliciting more negative interactions. Another possibility is perceptual or reporting bias, whereby individuals with more negative affect interpret benign interpersonal exchanges as “hassling.” A third explanation involves confounding from both observed factors (e.g., adverse childhood experiences, occupational environments) and unobserved traits that jointly shape exposure to negative social ties and biological aging. To address these sources of endogeneity and selection, we implement multiple strategies. First, we adjust for 3-year retrospective Charlson comorbidity index from the linked EHR data to reduce concerns about reverse causation, and we further examine whether the number of hasslers is significantly associated with self-reported health in a follow-up survey even after controlling for baseline health. Second, we incorporate occupation and psychosocial covariates to account for shared vulnerabilities and potential perceptual biases in reporting negative social interactions. Finally, we conduct sensitivity analyses to assess the magnitude of unobserved confounding required to explain away the estimated associations.

## Results

**Table S1** in the SI provides summary statistics for demographic characteristics and the key variables in our analytic sample (N = 2,345). Participants range widely in their chronological age, from 18 to 103 years (mean = 46.24, SD = 18.10), allowing us to explore epigenetic aging across a broad age span. We employ two complementary epigenetic aging measures: age-accelerated GrimAge2 (GrimAge2), which captures cumulative biological aging relative to chronological age, and DunedinPACE (PACE), which captures the rate of biological aging. Figure 1 illustrates that both GrimAge2 and PACE exhibit substantial variability across all chronological age groups. On average, older adults tend to display faster aging rates (PACE), but we also see individuals in younger age groups who show high epigenetic age acceleration (GrimAge2), indicating that some are aging faster biologically relative to what would be expected given their chronological age in the study population.

**Figure 1.**
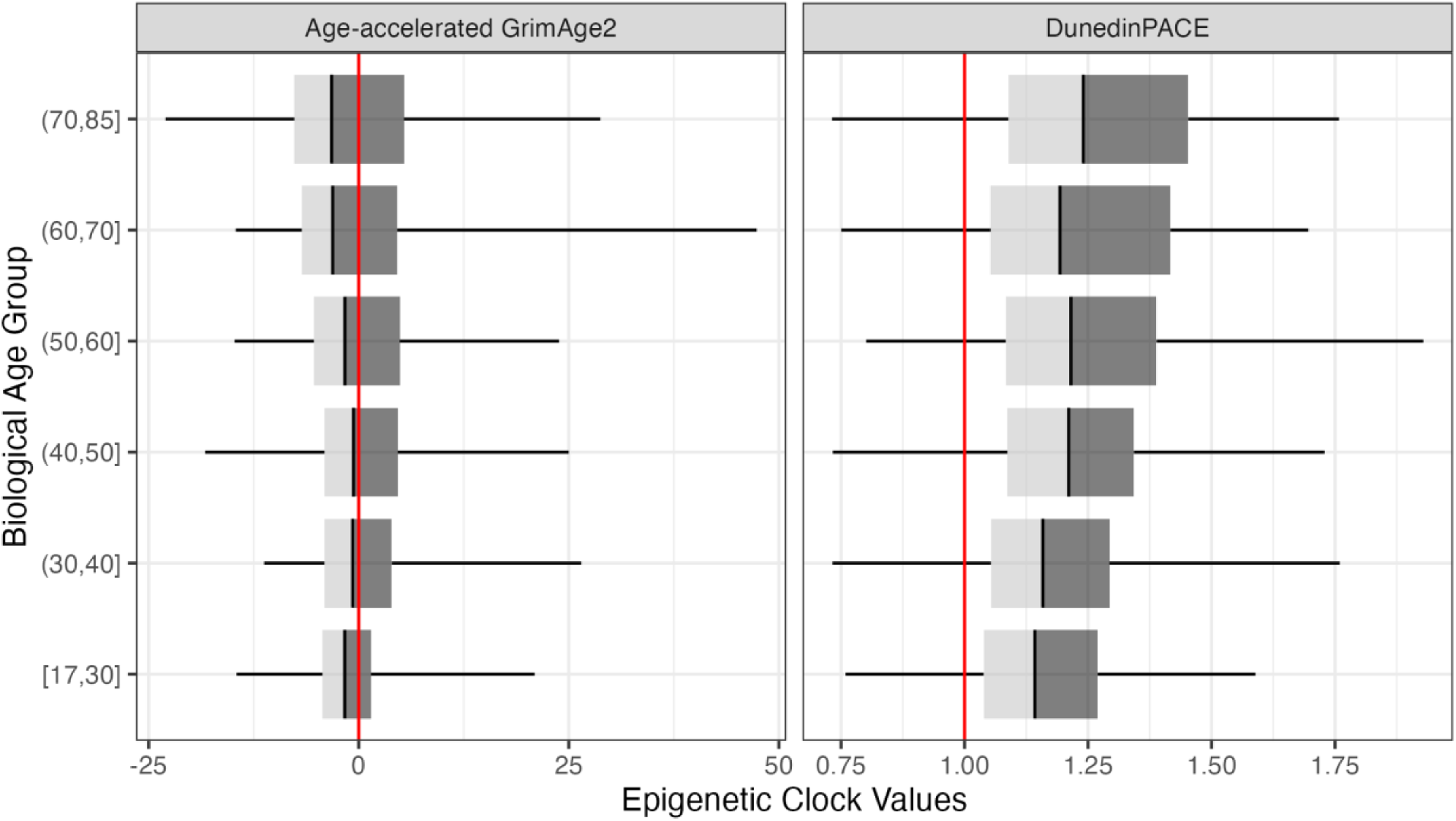
Distribution of Age-Accelerated GrimAge2 and DunedinPACE across chronological age groups. This figure presents the distribution of Age-Accelerated GrimAge2 (left panel) and DunedinPACE (right panel) across different chronological age groups. Each box plot displays the median (black line), interquartile range (dark gray box), and overall data range (whiskers). The red vertical lines indicate the reference points: 0 for Age-Accelerated GrimAge2 (indicating no deviation from expected aging) and 1.0 for DunedinPACE (representing the population average pace of aging).

### How many hasslers are present in egonetworks?

We define “hasslers” as network members whom respondents reported as “often” hassling them, causing them problems, or making life difficult, rather than selecting “never”, “rarely,” or “occasionally.” Table S2 presents the distribution of network size and the prevalence of hasslers within these networks. The average network size was 5.07 (SD = 2.46), with a maximum size of 25. Social isolation was rare, with only 1.3% of respondents reporting no network members, while 8.6% listed more than 10 names. On average, respondents identified 8.1% of their network members as hasslers. The overall proportion of hasslers remained relatively stable across different network sizes, although it tended to be higher among individuals with three or more members in their networks. Table S3 reports the distribution of hasslers. The mean size of the hassler network was 0.43 (SD = 0.85). Among 2,685 respondents, 28.8% reported having at least one hassler in their network, and about 10% indicated having two hasslers or more. These findings suggest that persistently negative ties are not rare among close social ties.

### Who is more likely to have hasslers in their egonetworks?

To investigate how demographic, occupational, psychosocial, and health-related characteristics predict exposure to negative social ties, we estimated survey-weighted zero-inflated Poisson regression models, which enable us to identify predictors of both having zero hasslers in one’s network (Panel A) and the expected number of hasslers (Panel B). Figure 2 plots the average marginal effects from these models (also see Table S4). Women had a significantly lower probability of having zero hasslers than men and, correspondingly, had higher expected counts of hasslers. Education, age, race, marital status, health insurance status, and life-time multiple morbidity showed insignificant associations in the multivariate models at p < 0.05. There are no significant differences across different occupation categories, although unemployed respondents in general tended to have more hasslers than those who were employed.

**Figure 2.**
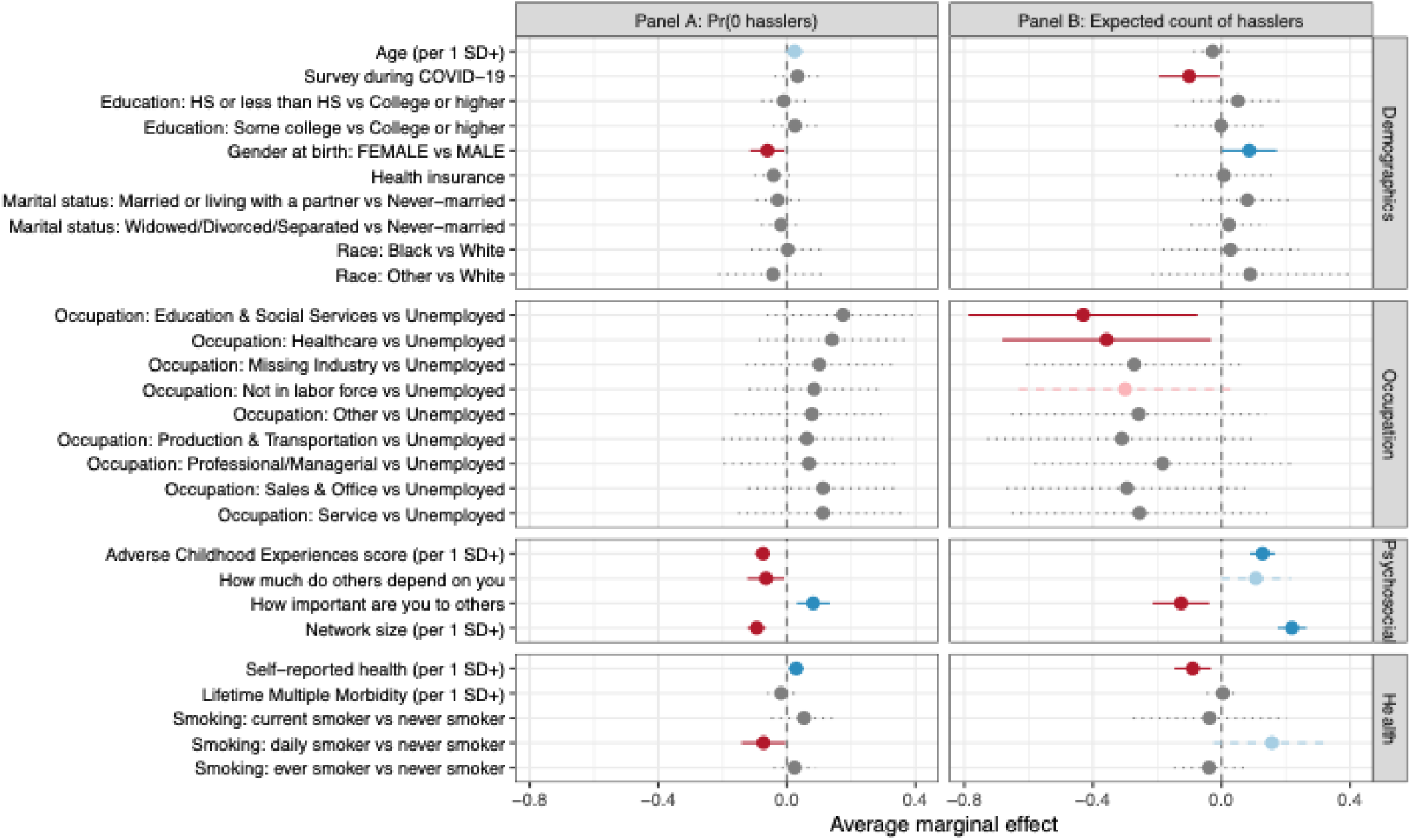
Determinants of Hasslers in Ego Networks from Zero-Inflated Poisson Models. Average marginal effects from zero-inflated Poisson regression models predicting (Panel A) the probability of having zero hasslers and (Panel B) the expected count of hasslers in respondents’ ego networks. Estimates are adjusted using survey weights and reflect changes in the outcome associated with a one–standard deviation increase in continuous predictors or contrasts between categorical groups, with 95% confidence intervals.

Psychosocial factors emerged as important predictors of hassler presence. The result from ACEs suggests that individuals with more adverse childhood experiences were more likely to have hasslers in their networks. A larger network size was associated with both a lower probability of having zero hasslers and a higher expected number of hasslers. Notably, respondents who felt that others depended on them were more likely to report a greater number of hasslers, whereas those who viewed themselves as important to others tended to exhibit the opposite pattern. In addition, health factors and behaviors predicted hassler connections. Better self-reported health was associated with both a higher likelihood of having zero hasslers and lower expected hassler counts. Daily smokers, compared with never smokers, showed a lower probability of having zero hasslers and higher expected counts. These results suggest that hassler exposure is not random but clustered around individuals with greater psychosocial and health vulnerabilities, underscoring the importance of examining whether such network strain contributes to accelerated biological aging.

### To what extent does exposure to hasslers through ego networks predict accelerated biological aging?

We examine associations between having hasslers and accelerated biological aging (AgeAccelGrim2) and the rate of aging (PACE) by estimating a series of OLS regression models (Models 1–6 in Table 1, also see Table S5 for the full regression coefficients) while controlling for demographic confounders (i.e., age, gender, education, marital status, and related technical background variables). Models 1 and 4 focus on the count of hasslers in networks. Individuals reporting more hasslers exhibit meaningful differences in both the rate and acceleration of biological aging. For PACE, each additional hassler is associated with a 0.015 unit increase (Model 1), corresponding to a 1.5% faster pace of aging. This means that rather than aging 1.00 biological year per calendar year, individuals with one more hassler age approximately 1.015 biological years annually. Although this yearly increase is modest, it accumulates over time: over a 10-year period, this faster rate results in about 1.8 extra months of biological aging for those with an additional hassler. For AgeAccelGrim2, which measures accelerated biological relative to chronological age, each additional hassler is associated with a nine-month increase (Model 4), indicating that individuals with one more hassler are, on average, about 9–10 months (0.782 * 12 months = 9.4) biologically older at the time of measurement than their peers of the same chronological age. To gauge the magnitude of these associations, we standardized both the independent and dependent variables in the regression models. The results indicate substantial associations: a one–standard deviation increase in the number of hasslers corresponds to a 0.065 SD increase in PACE and a 0.087 SD increase in AgeAccelGrim2. When benchmarked against the established effects of smoking—a major behavioral aging risk factor—these effect sizes correspond to approximately 17% of the PACE difference and 13% of the AgeAccelGrim2 difference between nonsmokers and smokers (0.382 SD for PACE; 0.657 SD for AgeAccelGrim2).

**Table 1.** Associations between Negative Social Ties and Epigenetic Aging Clocks.

|  | PACE |  |  | AgeAccelGrim2 |  |  |
| --- | --- | --- | --- | --- | --- | --- |
|  | Model1 | Model2 | Model3 | Model4 | Model5 | Model6 |
| Number of Hasslers | 0.0152**<br>(0.00382) |  |  | 0.782**<br>(0.172) |  |  |
| Presence of any hassler |  | 0.0259**<br>(0.00732) |  |  | 1.252**<br>(0.290) |  |
| N Hasslers (ref = 0) |  |  |  |  |  |  |
| 1 |  |  | 0.0110<br>(0.00782) |  |  | 0.707*<br>(0.282) |
| 2 |  |  | 0.0681**<br>(0.0157) |  |  | 2.339**<br>(0.340) |
| 3 |  |  | 0.0444<br>(0.0362) |  |  | 1.721<br>(1.159) |
| 4 |  |  | 0.0961+<br>(0.0523) |  |  | 4.494**<br>(1.076) |
| 5+ |  |  | -0.0126<br>(0.0222) |  |  | 2.615<br>(1.825) |
| Observations | 2345 | 2345 | 2345 | 2345 | 2345 | 2345 |
| Controls |  |  |  |  |  |  |
| Individual controls | V | V | V | V | V | V |
| Network size dummies | V | V | V | V | V | V |

Note. Coefficients represent the average marginal effects of negative social ties on biological aging outcomes, with standard errors in parentheses. Individual controls include age, gender, race/ethnicity, education, and marital status. Network size dummies account for differences in social network size, ensuring that the relative impact of negative ties is not confounded by absolute network size. Statistical significance is denoted as follows: + $p < .10$ , \* $p < .05$ , \*\* $p < .01$ (two-tailed tests).

We next consider the presence of any hasslers in Models 2 and 5, which show that presence of hasslers is linked to elevated aging scores (a 2.6% faster pace of aging and 15 months of accelerated biological aging). Lastly, Models 3 and 6 explore potential non-linear associations by estimating separate indicators for the number of hasslers. The results for PACE (Model 3) show that only individuals reporting two hasslers exhibit a statistically significant increase in biological aging, whereas higher counts display positive but imprecisely estimated effects. For AgeAccelGrim2 (Model 6), the association increases in magnitude as the number of hasslers rises, with particularly pronounced effects for those reporting two or more hasslers. Although the pattern is not perfectly monotonic, the results suggest that greater exposure corresponds to larger increases in epigenetic aging. Table S6 reports three alternative operationalizations of negative ties (i.e., the relative number of hasslers, categorical measures of the proportion of hasslers, and the mean frequency of hassling) and shows that the associations are robust across all measurement strategies.

### Which kinds of negative social ties matter most for accelerated biological aging, and why?

Not all hasslers might be the same. For example, the meaning of a hassler who is also a spouse might be different from the meaning of hasslers who are kin or non-kin. **Figure S1** shows the total number of hasslers by relationship type and the proportion of hasslers in each relationship category. Hassler prevalence varied considerably across relationship types. Among partner/spouse relationships, approximately 8.5% of spouses and 8.7% of partners were classified as hasslers. Within kin relationships, parents (9.8%) and children (9.7%) showed the highest proportions of hasslers, while grandparents (5.2%) and grandchildren (5.8%) had the lowest. Sibling and other kinship relationships fell in the middle range (both 7.5%). Non-kin relationships showed the widest variation in hassler prevalence. The “other” category had an exceptionally high proportion (40.5%) of hasslers, though this likely reflects a small group of heterogeneous relationships. Coworkers (11.3%) and roommates (10.4%) had notably higher hassler proportions compared to other non-kin ties. Friends, despite being a common relationship type, had a relatively low proportion (3.5%) of hasslers, as did church members (3.4%) and healthcare providers (4.1%). Neighbors showed a moderate proportion (6.5%). The substantial variation in hassler prevalence across relationship types implies that ties that involve obligation, shared space, or interdependence (e.g., parents, children, coworkers, roommates) appear more likely to produce hasslers, whereas voluntary and self-selected ties (e.g., friends, church members, healthcare providers) tend to generate fewer.

Given this substantial variation across relationship types, the next step is to determine whether the biological consequences of hasslers are equally heterogeneous. Table 2 presents the associations between spouse, kin, and non-kin hasslers and two epigenetic aging measures. The biological impact of hasslers varied substantially by relationship type. For PACE, kin hasslers showed the strongest associations with accelerated pace of aging. The presence of any kin hassler was associated with a 0.0264 year increase in PACE (p < .01), and each standard deviation increase in the number of kin hasslers was associated with a 0.00675 year increase (p < .05). In contrast, spouse hasslers and non-kin hasslers showed no significant associations with PACE, though coefficients were positive. For AgeAccelGrim2, both kin and non-kin hasslers showed significant associations: the presence of any kin hassler was associated with a 1.125 year increase in AgeAccelGrim2 (p < .01), while the presence of any non-kin hassler was associated with a 0.779 year increase (p < .05). When examining the standardized number of hasslers, both kin (0.352, p < .05) and non-kin hasslers (0.341, p < .05) showed similar magnitudes of associations. Spouse hasslers again showed no significant associations with AgeAccelGrim2, despite positive point estimates.

**Table 2.** Associations Between Spouse, Kin, and Non-Kin Hasslers and Epigenetic Aging Clocks.

|  | PACE |  | AgeAccelGrim2 |  |
| --- | --- | --- | --- | --- |
|  | Model1 | Model2 | Model3 | Model4 |
| Presence of any spouse hassler | 0.00772<br>(0.0162) |  | 0.852<br>(0.596) |  |
| Presence of any kin hassler | 0.0264**<br>(0.00869) |  | 1.125**<br>(0.329) |  |
| Presence of any non-kin hassler | 0.0150<br>(0.00903) |  | 0.779*<br>(0.336) |  |
| Standardized number of spouse hasslers |  | 0.00227<br>(0.00348) |  | 0.178<br>(0.113) |
| Standardized number of kin hasslers |  | 0.00675*<br>(0.00289) |  | 0.352*<br>(0.132) |
| Standardized number of non-kin hasslers |  | 0.00512<br>(0.00327) |  | 0.341*<br>(0.155) |
| Observations | 2346 | 2346 | 2346 | 2346 |

Note. Coefficients represent the average marginal effects of negative social ties on biological aging outcomes, with standard errors in parentheses. Individual controls include age, gender, race/ethnicity, education, and marital status. Network size dummies are included to account for variation in social network size, ensuring that the effects of negative ties are not confounded by absolute network size. Statistical significance is denoted as follows: \* $p < .05$ , \*\* $p < .01$ (two-tailed tests).

To further understand these heterogeneities, we assess relational characteristics of hasslers by examining network members’ tie strength, their degree centrality in ego’s network (i.e., how connected the alter is to other network members), and tie multiplexity (i.e., the extent to which ego has connections to network members through multiple relationship types across five different social support and exchange dimensions). Figure 3 presents survey-weighted comparisons of these network metrics between hasslers and non-hasslers across partner, kin, and non-kin relationship categories. Tie strength patterns revealed a consistent finding across all relationship types: hasslers maintained significantly weaker ties with egos than non-hasslers did. For alters’ network positions, non-hasslers held significantly more central positions within ego networks relative to hasslers. Yet within kin ties, the centrality of hasslers and non-hasslers did not differ significantly. When it comes to tie multiplexity, non-kin and kin hasslers had less multiplex ties than their non-hassler counterparts, indicating more uni-dimensional connections. Spouse hasslers showed a slight reduction in multiplexity relative to spouse non-hasslers, but the difference was not significant.

**Figure 3.**
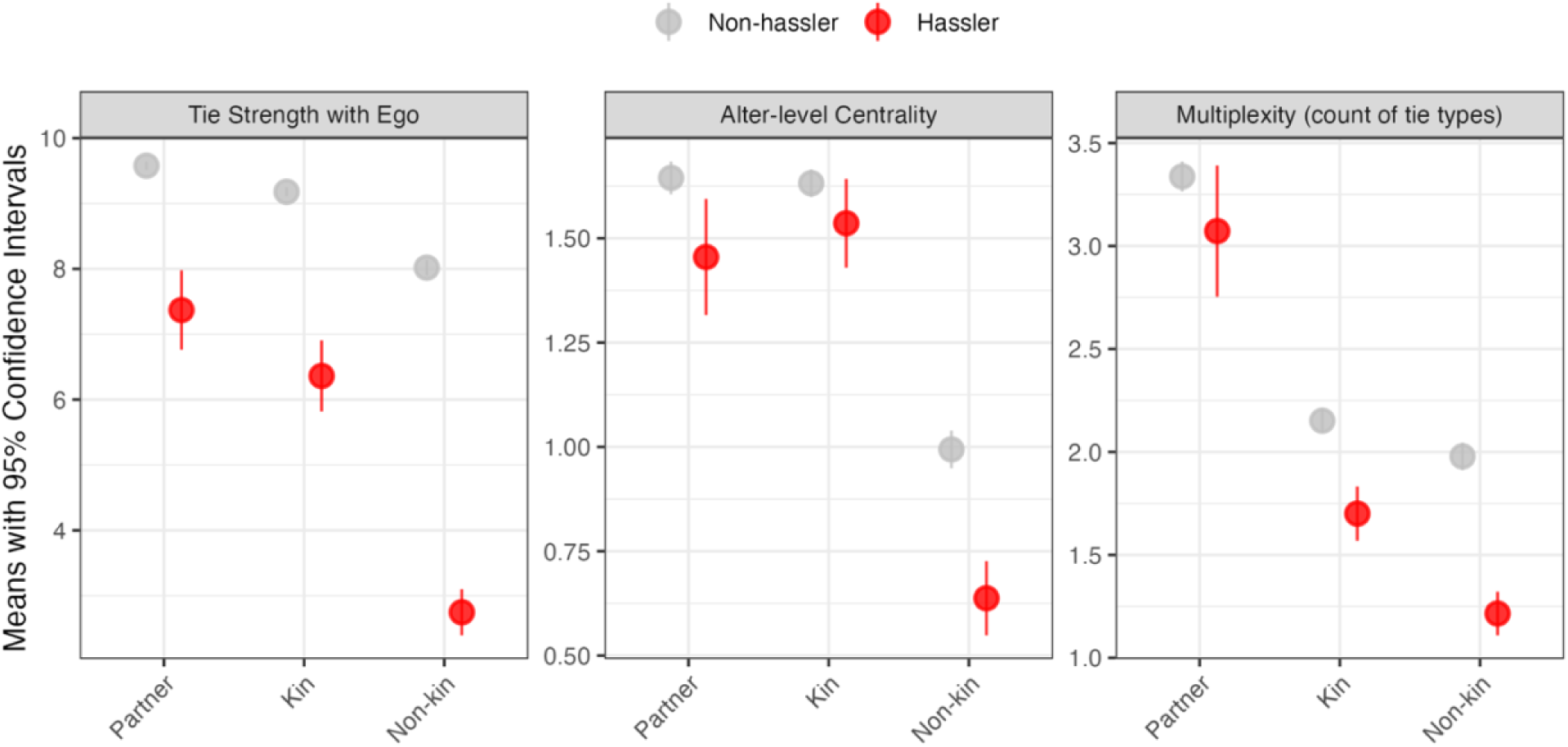
Differences in Tie Strength, Alter-level Centrality, and Multiplexity Between Hasslers and Non-hasslers Across Relationship Categories. Mean values (with 83% confidence intervals) of three alter-level characteristics for partners, kin, and non-kin, shown separately for hasslers and non-hasslers. *Tie strength* is measured on a 1–10 scale indicating the perceived strength of ego–alter relationships. *Alter-level centrality* reflects each alter’s average perceived closeness to all other alters in the ego network (0 = “don’t know each other,” 1 = “not very close,” 2 = “sort of close,” 3 = “very close”). *Multiplexity* is measured as the count (0–5) of relationship roles the alter occupies, including confidante, health discussant, health regulator, regular companion, and hassler. Across all relationship categories, hasslers tend to exhibit weaker ties to ego, lower centrality, and reduced multiplexity compared with non-hasslers, with the largest differences observed among non-kin alters.

These network patterns help explain why different types of hasslers show distinct associations with biological aging. Kin hasslers, who are more embedded and harder to avoid, exhibit the strongest and most consistent links to accelerated aging, especially for PACE, likely because their conflicts are both enduring and structurally inescapable. Non-kin hasslers, though weakly tied and peripheral, still predict faster aging on the more mortality-sensitive AgeAccelGrim2 clock, suggesting that even superficial strain may impose cumulative physiological burdens. Spouse hasslers show no significant associations, possibly because spousal ties mix negative and positive exchanges.

### Does the negative association of hasslers with health extend beyond biological aging?

Next, we assess whether the negative associations of hasslers are unique to biological aging or extend to broader health outcomes. Demonstrating similar patterns across a range of health outcomes strengthens the validity of our key findings on biological aging and is suggestive of wide-ranging pathogenic implications of molecular aging processes. Table 3 (see also Tables S7–S8 for full regression coefficients) shows that the number of hasslers in one’s network is consistently associated with worse health across multiple domains, even after adjusting for demographic characteristics and network size. Each additional hassler corresponds to significantly poorer self-rated general health, mental health, and physical health, as well as substantially higher anxiety severity and depression severity. These findings indicate that strained or conflictual social interactions are strongly linked to both psychological distress and diminished subjective well-being. The associations extend to objective physiological and clinical indicators: a greater number of hasslers predicts a higher epigenetic inflammation score, increased multimorbidity, a higher waist-to-hip ratio, and elevated BMI. It is important to note that the proportion of hasslers does not significantly affect height, which serves as a useful control because height is a stable biological trait unlikely to be influenced by social relationships in adulthood; thus, the absence of a significant association for height strengthens confidence that the observed associations with other health outcomes reflect genuine associations rather than methodological artifacts. These results underscore the broad and multifaceted impact of relational stressors on health trajectories.

**Table 3.**
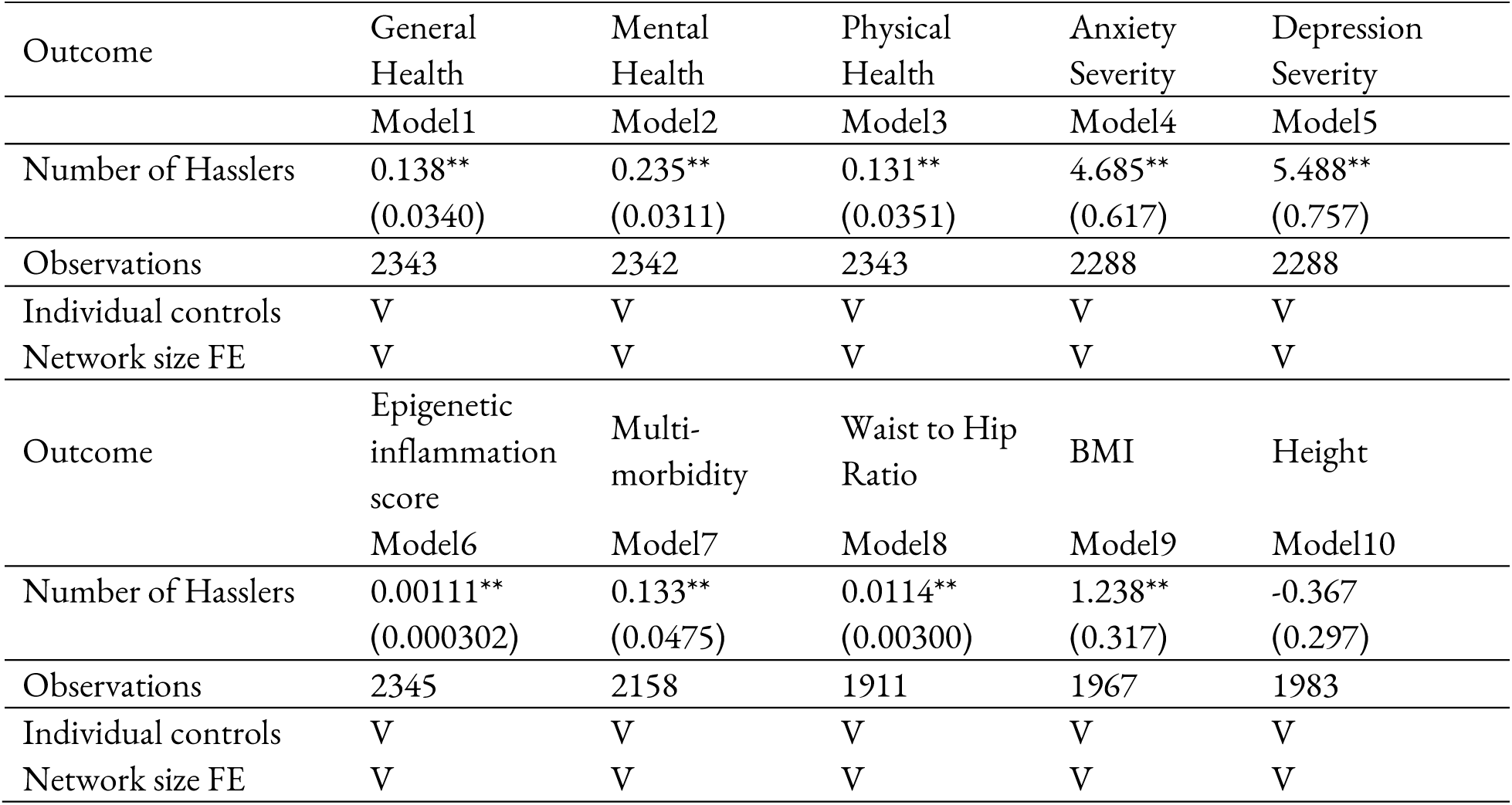
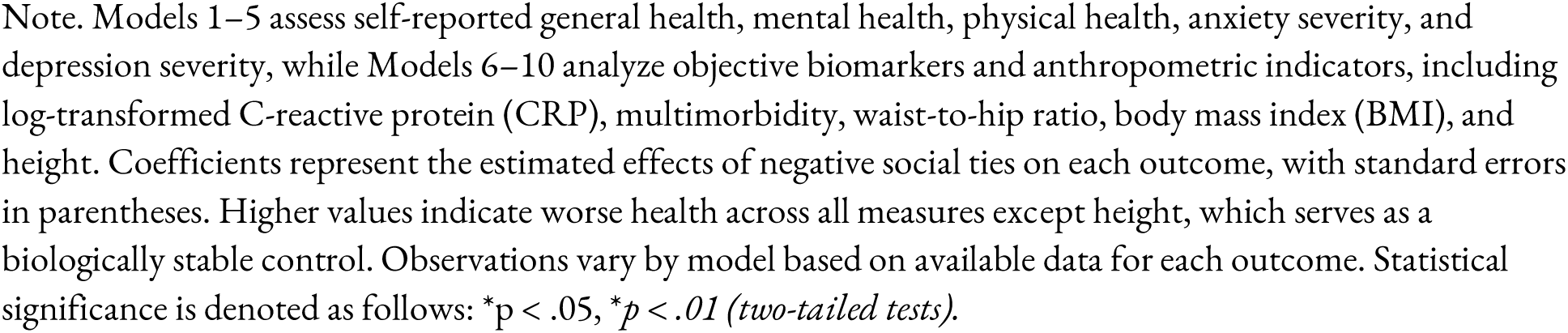
Associations between Negative Social Ties and Other Health Outcomes.

### To what extent is the association explained by confounding, selection, or reverse causality?

To evaluate the robustness of our findings, we conducted a series of sensitivity analyses that incorporated potential confounders of the relationship between negative social ties and biological aging. **Table 4** reports the average marginal effects of the number of hasslers on PACE (Models 1–8) and GrimAge2 (Models 9–16) as successive sets of controls are added to the baseline models (Models 1 and 9). In the PACE models, the number of hasslers remained significantly associated with faster biological aging after adjusting for insurance status and the timing of the COVID-19 pandemic. The association was attenuated but remained significant after further adjustment for the pre-survey Charlson comorbidity index (Model 3) and the lifetime multimorbidity index (Model 4), providing evidence against the health selection hypothesis. In Model 5, we accounted for two measures of affective orientation toward others to assess whether reports of “hasslers” primarily reflected respondents’ own dispositional tendencies rather than characteristics of their network alters. These controls did not meaningfully alter the magnitude or significance of the association, suggesting that the results are not artifacts of negative self- or other-perception. We next examined whether occupational and industry differences might confound the observed association, given that certain types of work expose individuals to heightened interpersonal strain and physiological stress. Model 6 shows that adjusting for occupation and industry did not substantially alter the estimated effect of hasslers on biological aging. In Models 7 and 8, the association attenuated but remained significant after adding controls for smoking (0.0110, p < 0.01) and adverse childhood experiences (0.0119, p < 0.01), respectively. Parallel patterns emerged in the GrimAge2 models. The initial association (0.782, p < 0.01) decreased to 0.498 (p < 0.01) after adjusting for smoking, suggesting partial mediation. Overall, the results show that although behaviors such as smoking explain part of the link, the association between negative social ties and accelerated aging remains strong, indicating an independent and enduring effect of social stress.

**Table 4.** Sensitivity Analysis of The Impact of Negative Social Ties on Epigenetic Aging Clocks.

| PACE |  |  |  |  |  |  |  |  |
| --- | --- | --- | --- | --- | --- | --- | --- | --- |
|  | Model1 | Model2 | Model3 | Model4 | Model5 | Model6 | Model7 | Model8 |
| Number of Hasslers | 0.0152**<br>(0.00382) | 0.0149**<br>(0.00388) | 0.0125**<br>(0.00445) | 0.0131**<br>(0.00396) | 0.0149**<br>(0.00378) | 0.0139**<br>(0.00375) | 0.0110**<br>(0.00348) | 0.0119**<br>(0.00404) |
| Observations | 2345 | 2339 | 2054 | 2345 | 2294 | 2345 | 2341 | 2345 |
| AgeAccelGrim2 |  |  |  |  |  |  |  |  |
|  | Model9 | Model10 | Model11 | Model12 | Model13 | Model14 | Model15 | Model16 |
| Number of Hasslers | 0.782**<br>(0.172) | 0.770**<br>(0.168) | 0.755**<br>(0.189) | 0.737**<br>(0.181) | 0.782**<br>(0.170) | 0.736**<br>(0.151) | 0.498**<br>(0.128) | 0.653**<br>(0.185) |
| Observations | 2345 | 2339 | 2054 | 2345 | 2294 | 2345 | 2341 | 2345 |
| Controls |  |  |  |  |  |  |  |  |
| Individual demographics | V | V | V | V | V | V | V | V |
| Network size FE | V | V | V | V | V | V | V | V |
| COVID-19 period |  | V |  |  |  |  |  |  |
| Health Insurance |  | V |  |  |  |  |  |  |
| Prior Charlson index |  |  | V |  |  |  |  |  |
| Medical visit (past 3yr) |  |  | V |  |  |  |  |  |
| Life-time multi-morbidity |  |  |  | V |  |  |  |  |
| Your importance to others |  |  |  |  | V |  |  |  |
| Others' dependance on you |  |  |  |  | V |  |  |  |
| Occupation |  |  |  |  |  | V |  |  |
| Smoking Status |  |  |  |  |  |  | V |  |
| ACEs |  |  |  |  |  |  |  | V |

Note. Models 1–8 estimate associations with PACE, and Models 9–16 estimate associations with AgeAccelGrim2. Coefficients represent the effects of the number of hasslers on each outcome, with standard errors in parentheses. All models adjust for demographic characteristics (age, gender, race/ethnicity, education, and marital status) and include additional robustness controls for survey period (COVID-19 exposure), health insurance status, Charlson multimorbidity index (from respondents' EHR records over the past 3 years), lifetime multi-morbidity index from the survey, psychosocial attitudinal measures, occupation, and smoking status (a known epigenetic confounder). Network size fixed effects are included to account for differences in the number of alters, ensuring that estimates are not confounded by absolute network size. Statistical significance: \* $p < .05$ , \*\* $p < .01$ (two-tailed tests).

Using longitudinal follow-up data collected between June and September 2022, we examined whether reverse causality could account for the associations observed at baseline. As shown in Table S9, baseline reports of hasslers significantly predicted worse follow-up health across all domains. Specifically, individuals with more hasslers reported poorer general health (Model 1: 0.145, *p* < .01), worse mental health (Model 3: 0.151, *p* < .01), and poorer physical health (Model 5: 0.231, *p* < .01). Importantly, although these associations were attenuated after adjusting for the corresponding baseline levels of each outcome, they remained statistically significant and substantively meaningful. Net of baseline health, baseline hasslers continued to predict declines in general health (Model 2: 0.106, *p* < .05), mental health (Model 4: 0.0795, *p* < .05), and physical health (Model 6: 0.132, *p* < .05).

This pattern supports temporal ordering from exposure to hasslers to subsequent health deterioration, offering strong evidence against reverse causality and reinforcing the interpretation that negative social ties may adversely shape later health.

To evaluate the robustness of the associations to potential unmeasured confounding, we conducted sensitivity analyses using the *konfound* tool (*30*). For PACE (see Table S10), the Robustness of Inference to Replacement (RIR) statistic indicated that 46.4% of the sample (1,088 cases) would need to be replaced by counterfactual cases with null effects to fully eliminate the association. The Impact Threshold for a Confounding Variable (ITCV) was 0.037, corresponding to required confounder-outcome and confounder-predictor correlations of 0.191. These values indicate that an unmeasured confounder would need to be stronger than nearly all observed covariates to overturn the effect; only leukocyte composition exhibits a comparable ITCV (0.034), and none of the sociodemographic controls come close. For AgeAccelGrim2, the RIR was similarly high—48.2% of the sample (1,129 cases)—and the ITCV was 0.039, implying threshold correlations of 0.198 with both the predictor and the outcome. Again, the required strength of an omitted confounder exceeds that of all observed covariates. These patterns demonstrate that the associations between the number of hasslers and both epigenetic aging measures are highly robust to omitted-variable bias.

## Discussion

Negative social ties have long been understood as a persistent element of human social life (*15*, *27*, *28*). Sociological and psychological research shows that these negative ties take many forms from everyday irritations and criticism to exclusion, hostility, denunciations, and even violence across a wide range of settings (*19*, *20*, *31*–*33*). Some negative ties dissolve when they destabilize broader network balance (*34*), some are leveraged to clarify status ambiguity or strengthen group cohesion (*35*), and others persist because of their ambivalent mix of support and strain (*19*, *20*, *36*). In everyday life, many individuals routinely encounter people who create problems or make life more difficult—what we refer to as *hasslers*. Their familiarity often leads people to normalize and endure them, which has resulted in surprisingly little attention to their long-term health implications. While supportive ties are well known to buffer age-related physiological decline (*12*, *37*–*39*), the absence of supportive ties is not equivalent to the presence of negative ones. By integrating comprehensive ego-centric network data with DNAm-based measures of biological age, this study provides novel evidence that negative social relationships operate as potent, chronic stressors capable of shaping epigenetic and physiological risk profiles across adulthood.

Prior work has documented that adverse social experiences ranging from loneliness to marital conflict can elevate allostatic load, increase inflammation, and shorten telomeres (*14*, *40*–*42*). Recent research using data on older adults from the Health and Retirement Study shows that both loneliness and social isolation, including the absence of friendships and marital relationships, are associated with epigenetic clocks measuring mortality risk and accelerated aging across multiple organ systems (*37*, *39*, *43*, *44*). However, few population-based studies have isolated the specific impacts of negative ties within networks of close relationships on biologically measured aging outcomes. Our data address this gap by linking directly measured negative ties to two state-of-the-art epigenetic clocks that capture both accelerated aging and the pace of aging. We find that respondents with more hasslers exhibit significantly greater epigenetic age acceleration, which is robust even after accounting for affective orientation, occupation, adverse childhood experiences, smoking, and prior co-morbidity in addition to demographic characteristics and network size. These results suggest that the “hasslers” in one’s social environment may constitute an overlooked but consequential biological risk factor. Each additional hassler is associated with roughly 1.5% faster aging and nine months of added biological age for individuals of the same chronological age, representing more than 13% and 17% of the estimated effect of smoking respectively. This is not a negligible impact, although it should be interpreted with caution and not taken as causal, as we discuss in the limitations below.

Our analyses show clearly that not all hasslers are the same. The nature of relationships and the network context substantially condition both the exposure to hasslers and their biological consequences. Ties characterized by obligation, shared space, or structural interdependence, such as parents, children, coworkers, or roommates, are more likely to be hasslers than voluntary, self-selected ties such as friends, church members, and neighbors. This heterogeneity extends to biology: kin hasslers are the most robust predictors of accelerated aging, while non-kin hasslers matter primarily for the mortality-sensitive AgeAccelGrim2 clock. In contrast, spouse hasslers show no significant associations with either measure. This pattern complicates existing work that emphasizes marital strain as a key driver of aging (*22*–*24*); our results suggest that spousal negativity may be buffered by the ambivalent mix of support and obligation within intimate partnerships, whereas kin conflicts, which are often emotionally salient yet hard to abandon, leave a clear imprint on aging.

Our network analyses help explain why. Kin hasslers tend to occupy still central positions within egos’ networks, but crucially, they tend to be *weaker in tie strength* and *less multiplex* than non-hassling kin ties, suggesting that these relationships are marked by obligation and structural embeddedness, but with reduced strength and fewer overlapping exchanges. While both embeddedness and multiplexity are usually protective because they offer support, monitoring, and multiple channels of reciprocity (*45*, *46*), when a tie is embedded *without* strong or multiplex connections–and it is *negative–*, the same structural features can trap individuals in repeated contacts with limited compensatory mechanisms. In other words, embeddedness that typically signals social protection can become a pathway for chronic stress when it is anchored in hasslers, turning “good structure” into biological risk.

Crucially, the detrimental influence of negative ties extends beyond epigenetic aging alone. When analyzing additional indicators—self-reported health, psychiatric symptoms, epigenetic inflammation score, waist-to-hip ratio, and BMI—those reporting a higher number of hasslers showed systematically worse health profiles across the board. Such convergence across molecular, anthropometric, clinical, and subjective outcomes strongly supports the idea that negative social relationships exact a broad physiological toll (*18*, *25*). These findings reinforce the concept of *allostatic load*, the cumulative “wear and tear” emerging from repeated physiological attempts to adapt to stress (*14*, *47*). Although positive social integration can reduce allostatic load, our study demonstrates the inverse: frequent exposure to hasslers may chronically activate stress-sensitive systems, fueling systemic inflammation, epigenetic dysregulation, and metabolic strain. These findings extend the literature by highlighting that negative social relationships are directly implicated in the same downstream physiological processes previously ascribed to major psychosocial and environmental stressors.

Negative social interactions could contribute to faster biological aging by putting chronic strain on the body’s hypothalamic-pituitary-adrenal (HPA) axis. When people experience frequent interpersonal tension, their bodies respond by activating stress-related systems, which trigger the release of hormones like cortisol and adrenaline (*48*). While these hormones help the body respond to immediate challenges, prolonged activation can take a toll on mental health, increasing feelings of anxiety and depression (*49*). At the same time, chronic social stress can fuel inflammation in the body. When stress is persistent, it activates molecular pathways that increase the production of inflammatory proteins, and over time, this repeated immune activation leads to higher levels of epigenetic inflammation scores. Our findings on the significant associations between the number of hasslers in close social ties and both mental health and inflammation markers provide indirect evidence to support these mechanisms.

Our findings reveal that exposure to negative social ties is not evenly distributed but it is instead patterned along lines of psychosocial and health vulnerability. In particular, women, daily smokers, those in poorer health, and those with greater histories of adverse childhood experiences (ACEs) were all more likely to have (more) hasslers in their networks. These inequalities have important implications for understanding how social environments may compound disadvantage across the life course (*50*, *51*). Individuals with greater psychosocial adversity, such as those with higher ACEs, appear not only to carry early-life scars but to inhabit adult relational contexts where difficult, demanding, or conflict-prone ties are more common. Poorer health and health-behavior risks show a similar pattern: those already at elevated physiological risk are simultaneously embedded in social networks that may amplify stress exposure. These patterns reflect a form of “relational inequality,” in which unequal exposure to negative relationships becomes a mechanism through which broader health disparities are reproduced (*52*).

Several limitations merit attention. First, because our analyses are cross-sectional, we cannot establish a definitive causal pathway between negative ties and biological aging; longitudinal or quasi-experimental designs are needed to clarify how hasslers emerge and whether they prospectively accelerate aging. We took multiple steps to assess alternative explanations, including adjustment for (a) self-perceptual bias (i.e., the possibility that reporting hasslers reflects respondents’ dispositions rather than objective network features), (b) occupational context that might jointly increase exposure to negative interactions and independently influence aging via job stress, schedules, or economic precarity, (c) adverse childhood experiences, (d) health selection using prior comorbidity and longitudinal data on self-reported health, and (e) smoking behavior. These checks increase confidence in the robustness of our associations, but they should not constitute causal estimates.

Second, hasslers are measured by self-report and may be vulnerable to recall or appraisal biases. We partially address this concern by controlling for affective orientation toward others. Still, future studies using repeated measures or alter-reports would strengthen inference. Third, our results may be affected by mortality selection. Individuals with poorer health, lower support, or chronic stress are more likely to die before data collection, yielding an analytic sample that may be healthier and more socially advantaged than the population at risk. If the highest-risk individuals are disproportionately missing, our estimates likely understate the true impact of negative ties. However, age-specific analyses indicate that associations are significant among young adults (18–30) for PACE and broadly similar across age groups for AgeAccelGrim2 (Table S11), which suggests that mortality selection alone is unlikely to account for the findings. Nevertheless, because we cannot directly model mortality selection, these limitations continue to caution against causal interpretation.

Finally, while we conceptualize hassling as one dimension of negative interaction within ego networks, our survey includes only a single explicit indicator of negative relational behavior. We therefore cannot assess whether hassling co-occurs with other forms of negative ties—such as hostility, coercion, chronic criticism, or gaslighting—or whether different negative-tie subtypes have distinct biological consequences. Given longstanding arguments that negative exchanges take multiple forms within networks, future work would benefit from richer measurement of negative ties. Such data would allow researchers to differentiate various kinds of relational strain, test whether hasslers represent a unique subtype or a broader negative-tie pattern, and more precisely locate hassling within the interpersonal processes that shape health and aging.

Taken together, our findings indicate that negative social relationships may function as powerful chronic stressors, accelerating age-related biological processes through multiple, interlinked pathways. By systematically mapping these relationships onto advanced epigenetic measures and additional health markers, this study highlights that the “dark side” of social connections can wear down physiological resilience and hasten aging and the development of multiple morbidities. Future strategies to mitigate relationship strain, alongside bolstering positive support, could represent a crucial avenue to promote healthier aging and reduce the burden of age-related disease. Rather than focusing solely on loneliness or isolation, these results underscore the need to recognize and address the multifaceted nature of social adversity in shaping our biological and aging trajectories.

## Data and Methods

This study utilizes data from the Person-to-Person Health Interview Study (P2P), a statewide omnibus health survey conducted in Indiana (*53*). The P2P provides representative estimates for Indiana’s adult population (aged 18 years and older) on various health, social, and behavioral measures. Employing a stratified probability sampling design similar to the General Social Survey, the study drew participants from household addresses in both rural and urban counties across Indiana. Sampling was executed by the National Opinion Research Center at the University of Chicago, with intentional oversampling in two economically disadvantaged rural counties. Trained interviewers administered structured surveys using standardized protocols and collected anthropometric measurements. At the end of each interview, participants were invited to provide saliva samples for epigenetic analysis. DNA extraction and quality assessment occurred within six weeks of collection at the Indiana University Genetics Biobank using the Chemagic 360 magnetic bead-based high-throughput extraction system. Data collection spanned from October 2018 to July 2021, with 2,685 individuals participating (overall response rate of approximately 30%). Of these, 87.4% (N = 2,347) consented to saliva collection. An additional 10 respondents were excluded due to missing data on key control variables, predictors, or outcomes. The final analytic sample included 2,345 participants for analyses predicting biological aging clocks, though the exact sample size varied slightly across regression models depending on the specific control variables and outcome measures included. For a subset of the analytic sample (N = 1,271), we conducted a follow-up survey between June and September 2022 that collected self-reported health measures, although longitudinal epigenetic aging clocks were not available for this wave. The Institutional Review Board at Indiana University approved all study procedures. Informed consent was obtained from each participant, and the confidentiality of data was rigorously maintained throughout the study.

### Egocentric Network Module

A central aspect of the P2P survey is its ego-centric network module, which captures respondents’ (“egos”) significant social connections (“alters”). Participants were asked to complete a personal social network protocol following standard methods of egocentric network research (*54*). Initially, respondents were asked via “name generators” to list individuals with whom they have interacted over the past six months. Here, the number of individual respondents could name was unrestricted, avoiding potential right censoring common in similar ego-centric network surveys. The survey employed five name generators from the PhenX Social Network Battery (*55*) to elicit different types of relationships: confidantes (people with whom respondents discussed personal problems), health discussants (individuals respondents discussed their health with or could rely on for help regarding physical or emotional issues), health regulators (those frequently discussing and influencing respondents’ mental and physical health behaviors), regular companions (people respondents regularly spent free or leisure time with), and hasslers (individuals causing respondents stress or difficulty). Following the elicitation of names, respondents completed “name interpreter” questions that detailed each alter’s characteristics and relationship attributes.

To identify negative ties, we ask each ego to assess each alter’s potential for interpersonal stress or conflict in the following question: “How often has (alter) hassled you, caused problems, or made life difficult?” Here, we define “difficult ties” (i.e., hasslers) as alters whom respondents identified as “often” causing problems or difficulties, rather than “occasionally”, “rarely” or “never.” We do not consider “occasional hassling” as hasslers since this likely captures normative, supportive informal social control and should not be interpreted as negative. Our primary measures in this study are the count of hasslers within each respondent’s network. We present results assuming zero hasslers when network size is zero, though the findings remain virtually unchanged if we drop these cases, as very few individuals (N = 35, 1.3%) report a network size of zero. To examine the heterogeneous influence of spouse, kin, and non-kin hasslers, we first classify all alters into three relationship categories: spouse (spouse or partner); kin (parent, sibling, child, grandparent, grandchild, or other relative); and non-kin (friend, coworker, neighbor, roommate, church member, health provider, or other). Because alters may occupy multiple relational roles across the network, we allow an alter to contribute to more than one category when appropriate. For each respondent, we then count the number of hasslers within each category. To assess robustness and comparability across relationship types, we examine both binary indicators of whether any spouse/kin/non-kin hassler is present and standardized counts of hasslers within each category.

To characterize the relational context of hassling ties, we examine several features of each alter who engages in hassling. First, we measure tie strength using respondents’ ratings of how strong their relationship is with each alter on a 1–10 scale (10 = strongest). Second, we assess each alter’s degree centrality within the ego network, capturing how connected that alter is to other network members. Degree centrality is computed from pairwise closeness ratings in which respondents indicate how close each pair of alters is to one another (0 = don’t know each other, 1 = not very close, 2 = sort of close, 3 = very close). For each alter, we take the mean closeness score across all other alters. Finally, we measure tie multiplexity, defined as the number of distinct social support or exchange domains (out of five possible types) through which ego and alter are connected. This count reflects the extent to which the relationship spans multiple functional dimensions of social involvement.

### Saliva Collection and Epigenetic Data Processing

DNA methylation (DNAm) was assessed from saliva samples using the Infinium Methylation EPIC BeadChip. To ensure unbiased results, samples were randomized across plates based on demographic variables (age, sex, and race/ethnicity), and 40 pairs of blinded duplicate samples were included. Data preprocessing and quality control were conducted using the *minfi* package in **R**, with probes and samples failing the detection P-value threshold excluded, along with samples demonstrating sex mismatches. This study focuses on two advanced DNAm clocks—age-accelerated GrimAge2 and DunedinPACE—both of which demonstrate superior predictive validity for morbidity and mortality compared to earlier-generation epigenetic clocks. Age-accelerated GrimAge2 (AgeAccelGrim2) and DunedinPACE (PACE) offer complementary insights into biological aging by capturing both cumulative biological aging and the current rate of physiological aging respectively, providing a robust, multidimensional evaluation of aging processes and associated longevity risks.

GrimAge2 was developed using a two-stage procedure: first, plasma proteins and smoking pack-years were regressed on chronological age, sex, and CpG methylation levels to generate DNAm-derived biomarkers. These biomarkers were then used to predict time-to-death, adjusting for age, sex, and DNAm-smoking-pack-years (*6*). We use the age-adjusted version (i.e., age regressed out) of GrimAge2, which represents the residual difference between observed and predicted GrimAge2, with higher values indicating accelerated biological aging beyond what is expected for a given chronological age. Elevated AgeAccelGrim2 scores are associated with increased vulnerability to mortality and age-related diseases, making it a robust predictor of long-term health risks and physiological decline. In contrast, DunedinPACE quantifies the ongoing rate of biological aging rather than cumulative age acceleration. The creation of this measure also involved multiple phases (*5*). First, researchers tracked longitudinal changes in 19 biomarkers reflecting multiple organ systems, assessed at ages 26, 32, 38, and 45. They then used linear mixed-effects models to estimate the rate of change for each biomarker, which were combined to calculate individual pace of aging scores. These scores were scaled so that a value of 1.0 represents one biological year per chronological year (e.g., a score of 2.3 indicates aging at 2.3 biological years per chronological year). Finally, researchers applied elastic net regression to create an algorithm that predicts the 20-year pace of aging using a subset of DNAm markers measured at age 45. Thus, unlike GrimAge2, PACE provides a dynamic measure of aging speed, directly capturing the rate of cellular and physiological deterioration.

It is important to note that saliva contains both leukocytes and epithelial cells, which differ from the peripheral blood leukocyte mixtures used to develop the original GrimAge2 and DunedinPACE clocks. Because epithelial cells tend to exhibit methylation patterns associated with older or faster biological aging, saliva-based DNAm clocks can yield higher absolute levels than the blood-based reference distributions. However, salivary methylation estimates have been widely used for both PACE (*56*) and GrimAge2 (*57*), and their rank-ordering and associations with exposures remain stable when applied to saliva, even if absolute levels differ due to cell-type composition (*58*), which is not surprising given that immune cells account for more than 80% of the DNA present in saliva (*59*). Nevertheless, comparisons of our sample’s mean AgeAccelGrim2 or PACE values to the original reference populations should be interpreted cautiously. The mean AgeAccelGrim2 in our sample is -0.69 (SD = 7.04), indicating that participants, on average, closely match the GrimAge2 reference population for cumulative epigenetic aging, though considerable variation exists at the individual level (range: -22.99 to 47.36 years). By contrast, the mean PACE is 1.19 (SD = 0.19), above the benchmark rate of 1.0 established in the original Dunedin Study cohort, though it may be due to the nature of the saliva sample in our study.

### Other Health Outcomes

*Self-rated general health* was measured by asking “Would you say that, overall, your health is…” with response options: 1) Poor, 2) Fair, 3) Good, 4) Very Good, 5) Excellent. *Self-rated physical health* was measured by asking “Would you say that, in general, your mental health is…” with response options: 1) Poor, 2) Fair, 3) Good, 4) Very Good, 5) Excellent. *Self-rated mental health* was measured by asking “Would you say that, in general, your physical health is…” with response options: 1) Poor, 2) Fair, 3) Good, 4) Very Good, 5) Excellent. *Depression and anxiety severity* (range = 0 – 100) were measured using the Computerized Adaptive Test‒Mental Health (CAT-MH) (*60*, *61*), an efficient computerized adaptive test based on multidimensional item response theory. The CAT-MH contains validated measures for diagnostic screening and continuous measurement of targeted mental health disorders and is superior to brief assessments used in other studies.

*Multimorbidity* (range = 0 - 9) was measured by summing the total number of affirmative responses to a set of question asking whether respondents had ever been diagnosed by a physician with the following conditions: arthritis, asthma, cancer, emphysema, stroke, myocardial infarction, coronary heart disease, kidney disease, obesity, and diabetes. *Epigenetic inflammation score* is a measure derived from a large-scale epigenome-wide association study (EWAS) of circulating C-reactive protein (CRP), a widely used biomarker of systemic inflammation, which identified and replicated DNAm signatures at 58 CpG sites across two large and ethnically diverse cohorts (N > 12,000). These methylation markers were robustly associated with CRP levels and showed enrichment in genes linked to immune response and cardiometabolic risk, independent of known genetic predictors. By leveraging these validated methylation sites, we construct a composite inflammation score that reflects chronic inflammatory processes at the molecular level (*62*).

For anthropometric measurement, trained interviewers measured participants’ weight and height three times using a standardized protocol to ensure accuracy and reliability. Waist and hip circumferences were also measured three times using a flexible measuring tape at the narrowest part of the waist and the widest part of the hips, respectively. The final weight, height, waist, and hip values used in analyses were the averages of the three measurements. The *waist-to-hip ratio* was calculated as the average waist circumference divided by the average hip circumference. *Body Mass Index* (BMI) was computed as weight (kg) divided by height squared (m²). These anthropometric indicators were used to assess overall body composition and central adiposity in subsequent analyses.

### Controls

Consistent with standard practice for models regressing epigenetic clocks on predictors, we adjust for two technical covariates: batch effects and cell types (*63*, *64*). We control for the following socio-demographic characteristics across all models: age, gender at birth (men/women), race and ethnicity (non-Hispanic White / non-Hispanic Black / Other race), educational attainment (less than high school education or high school diploma or equivalent / some college or technical school/bachelor’s degree or higher), and marital status (never married/married or living with a partner / divorced or widowed or separated).

In the sensitivity analysis, we account for additional potential post-treatment confounders that might influence both social network characteristics and epigenetic aging to rigorously assess the robustness of our findings. First, the COVID-19 pandemic occurred while the P2P study was in the field and data collection was suspended for one year during the height of the pandemic. The COVID-19 pandemic might have induced differences in social network patterns (*65*) and/or affected epigenetic aging (*66*). As such, we adjust for a variable indicating whether the data was collected before or after the pandemic. Second, we adjust for health insurance status, which captures disparities in access to medical care and preventive services, as it may shape both biological aging and the structure of social relationships. Third, to account for underlying morbidity burden and reduce the risk of reverse health selection, we constructed a Charlson Comorbidity Index based on ICD-9/10 diagnoses recorded in the Indiana Network for Patient Care (INPC) electronic health record data (*67*). Using the comorbidity **R** package (*68*) and the original Charlson weighting scheme (*69*), we mapped all qualifying diagnoses occurring within the three years prior to respondents’ interview dates. Diagnoses were identified by linking all encounter-level information three years prior to survey dates, and applying an established severity hierarchy. We imputed zeros for patients who had at least one encounter but no qualifying diagnoses. Because individuals with limited healthcare contact may appear artificially “healthy,” we also constructed an indicator for any encounter in the prior three years and adjusted for it in all regression models to mitigate ascertainment bias. Fourth, to capture their affective orientation toward others, we use two survey items—*your importance to others* (“How important are you to others?”) and *others’ dependence on you* (“How much do other people depend on you?”)—from the mattering scale (*70*) and binarize them (coded 1 = “a lot,” and 0 = “somewhat,” “a little,” or “not at all”).

Fifth, we construct occupational categories using a multistep process that integrates structured industry checklists with open-ended occupational histories, and harmonized coding schemes. In the survey, respondents indicated whether they had ever worked in any of a predefined set of industries (i.e., farming, automotive, construction, manufacturing, railroad, forestry, electrical, beauty services, welding, firefighting). For respondents whose primary occupation was not captured by these industry options, they list any occupations in which you worked five or more years in the open-ended response format, which trained research assistants code into standardized occupational categories using the SOC 2018 scheme. We then merged the coded open-ended occupations with the structured industry indicators to produce a unified combined occupation label that preserves the most specific available information. Then, to facilitate analysis, we grouped the combined occupation labels into ten broader occupational groups: (1) Professional/Managerial (2) Healthcare (3) Education and Social Services, (4) Service (5) Sales and Office, (6) Production and Transportation (7) Other (e.g., arts, entertainment, media, and military-specific occupations), (8) Not in labor force, (9) Unemployed, and (10) Missing Industry (respondents with employment but insufficient information to classify). Sixth, we adjust for smoking status (never smoker / ever smoker / current smoker / daily smoker) to determine whether the association between social network characteristics and epigenetic clocks is behavioral rather than biological. Finally, we measure childhood experiences (ACEs), as prior literature has consistently linked early-life adversity with accelerated biological aging trajectories, using the 12-item index, which assesses potentially traumatic events that occur in childhood (0-17 years), such as experiencing violence, abuse, or neglect, witnessing violence in the home, or substance abuse in the home (*71*) (Cronbach’s alpha = 0.83).

### Analytic Strategy

Our analytic approach focuses on evaluating how negative social ties (hasslers) within individuals’ social networks relate to accelerated biological aging, as measured by two DNAm-based epigenetic clocks: age-accelerated GrimAge2 (AgeAccelGrim2) and PACE. The primary independent variable is the count of hasslers reported by each respondent. We employ OLS regression models to estimate associations between negative social ties and our biological aging outcomes. Our initial models include basic socio-demographic covariates—age, gender, race/ethnicity, education, and marital status—to adjust for fundamental demographic confounding. We also introduce dummy variables indicating network size categories to accounts for the possibility that respondents with larger networks may report a higher absolute count of hasslers, clarifying the independent contribution of network negativity above and beyond network size itself.

To further assess the robustness and validity of our results, we conduct a series of sensitivity analyses. In doing so, we explore alternative specifications of negative ties to assess potential nonlinearities or threshold effects in several complementary ways: We construct a binary indicator for whether any hassler is present in a respondent’s network; we categorize the number of hasslers using discrete dummy variables (0, 1, 2, 3, 4, 5+); we measure the proportion of network members who are hasslers and also create categorical versions of this measure (0%, 1–24%, 25–49%, 50–99%, 100%). These analyses examine whether biological aging accelerates disproportionately when negative ties surpass a critical threshold within an individual’s close relationships. In addition, we capture overall intensity by computing the mean hassling frequency across all alters (coded 3 = often, 2 = occasionally, 1 = rarely, 0 = never).

To address potential selection and endogeneity concerns, we sequentially introduce additional controls that could confound the observed relationship between negative social ties and biological aging. Moreover, as a negative control outcome, we test the association between baseline hasslers and adult height, for which no association is expected. In addition, using a subset of samples we followed up longitudinally, we estimate lagged dependent variable (LDV) models for self-rated health, regressing follow-up self-rated health on baseline hasslers while adjusting for baseline self-rated health and covariates. To quantify how strong unmeasured confounding would need to be to overturn our findings, we use the *konfound* Stata package to compute two sensitivity indices (*30*): the Impact Threshold for a Confounding Variable (ITCV) and the Robustness of Inference to Replacement (RIR). The ITCV quantifies the smallest correlation that a single omitted confounder would need to have with both the exposure and the outcome, conditional on the set of observed covariates, to reduce our estimated effect below a chosen threshold. Notably, results are substantively identical when using the unconditional version. The RIR expresses the minimum proportion of cases in the sample that would have to be hypothetically replaced by cases showing no effect of the exposure in order to invalidate the inference. Finally, we expand our analyses to other health outcomes, including self-rated health, mental health symptoms (anxiety and depression severity scores), inflammation, and anthropometric indicators (waist-to-hip ratio, BMI). This comprehensive assessment evaluates whether negative social relationships exert broad-spectrum health impacts beyond biological aging itself, reflecting chronic stress-induced multisystem physiological dysregulation.

We interpret and report results as average marginal effects (AMEs) to facilitate intuitive understanding and direct comparison across models. To ensure the generalizability and representativeness of our findings to the adult population of Indiana, all analyses incorporate survey weights provided by NORC. These weights correct for differential nonresponse rates and oversampling of specific demographic subgroups. Analyses are performed using Stata and **R** statistical software, and all analytic codes required to reproduce our results will be publicly shared upon publication.

## Data Availability

Replication code will be made publicly available upon publication of the paper.

## Supporting Information

**Table S1.** The descriptive statistics of individual characteristics and the key variables in the analytic sample.

|  | Mean | SD | Min | Max | N (=2345) |
| --- | --- | --- | --- | --- | --- |
| <u>Individual characteristics</u> |  |  |  |  |  |
| Age | 46.24 | 18.10 | 18 | 104 |  |
| Gender: Female | 0.52 | 0.50 | 0 | 1 |  |
| Race: White | 0.81 | 0.39 | 0 | 1 |  |
| Race: Black | 0.10 | 0.31 | 0 | 1 |  |
| Race: Hispanic | 0.09 | 0.28 | 0 | 1 |  |
| Education: HS or less than HS | 0.30 | 0.46 | 0 | 1 |  |
| Education: Some college | 0.38 | 0.49 | 0 | 1 |  |
| Education: College or higher | 0.32 | 0.46 | 0 | 1 |  |
| Marital Status: Never-married | 0.23 | 0.42 | 0 | 1 |  |
| Marital Status: Married or cohabiting | 0.57 | 0.49 | 0 | 1 |  |
| Marital Status: Widowed/Separated/Divorced | 0.20 | 0.40 | 0 | 1 |  |
| Occupation: Professional/Managerial | 0.06 | 0.23 | 0 | 1 |  |
| Occupation: Healthcare | 0.07 | 0.25 | 0 | 1 |  |
| Occupation: Education & Social Services | 0.05 | 0.21 | 0 | 1 |  |
| Occupation: Service | 0.06 | 0.23 | 0 | 1 |  |
| Occupation: Sales & Office | 0.07 | 0.26 | 0 | 1 |  |
| Occupation: Production & Transportation | 0.01 | 0.12 | 0 | 1 |  |
| Occupation: Other | 0.01 | 0.09 | 0 | 1 |  |
| Occupation: Not in labor force | 0.22 | 0.42 | 0 | 1 |  |
| Occupation: Unemployed | 0.04 | 0.19 | 0 | 1 |  |
| Occupation: Missing Industry | 0.42 | 0.49 | 0 | 1 |  |
| Smoking: Never smoker | 0.55 | 0.50 | 0 | 1 | 2341 |
| Smoking: Ever smoker | 0.22 | 0.42 | 0 | 1 | 2341 |
| Smoking: Current smoker | 0.06 | 0.24 | 0 | 1 | 2341 |
| Smoking: Daily smoker | 0.17 | 0.38 | 0 | 1 | 2341 |
| % Leukocyte cells | 0.71 | 0.19 | 0 | 0.98 |  |
| Batch Number (ref = 8615) | 0.15 | 0.36 | 0 | 1 |  |
| Batch 8732 | 0.18 | 0.39 | 0 | 1 |  |
| Batch 9054 | 0.24 | 0.43 | 0 | 1 |  |
| Batch 9109 | 0.22 | 0.41 | 0 | 1 |  |
| Batch 9213 | 0.15 | 0.36 | 0 | 1 |  |
| Batch 11277 | 0.01 | 0.12 | 0 | 1 |  |
| Batch 13762 | 0.04 | 0.20 | 0 | 1 |  |
| Interview during COVID-19 | 0.40 | 0.49 | 0 | 1 |  |
| Has Health Insurance | 0.92 | 0.28 | 0 | 1 | 2339 |
| Adverse childhood experiences (count) | 2.39 | 2.67 | 0 | 12 |  |
| How important are you to others (binary) | 0.68 | 0.47 | 0 | 1 | 2299 |
| How much do others depend on you (binary) | 0.65 | 0.48 | 0 | 1 | 2336 |
| <u>Independent variables</u> |  |  |  |  |  |
| Network Size: All | 5.29 | 2.95 | 0 | 25 |  |
| Network Size: Negative ties | 0.42 | 0.88 | 0 | 8 |  |
| % Hassler | 0.07 | 0.16 | 0 | 1 |  |
| Mean Hassle Frequency | 0.84 | 0.65 | 0 | 3 |  |
|  | 0.06 | 0.23 | 0 | 2 |  |
|  | 0.22 | 0.67 | 0 | 7 |  |
|  | 0.24 | 0.68 | 0 | 8 |  |
|  | 0.06 | 0.23 | 0 | 1 |  |
|  | 0.14 | 0.35 | 0 | 1 |  |
|  | 0.16 | 0.37 | 0 | 1 |  |
| <u>Outcome variables</u> |  |  |  |  |  |
| AgeAccelGrim2 | -0.69 | 7.04 | -23.00 | 47.37 |  |
| PACE | 1.19 | 0.19 | 0.73 | 1.93 |  |
| Self-reported General Health | 2.69 | 0.99 | 1 | 5 | 2343 |
| Self-reported Mental Health | 2.56 | 1.06 | 1 | 5 | 2342 |
| Self-reported Physical Health | 2.84 | 1.04 | 1 | 5 | 2343 |
| Epigenetic inflammation score | 0.06 | 0.03 | 0.00 | 0.17 |  |
| Charlson co-morbidity index (past 3yr) | 0.28 | 0.90 | 0 | 8 | 2054 |
| Any medical visit (past 3yr) | 0.71 | 0.45 | 0 | 1 | 2054 |
| Multi-morbidity index | 1.09 | 1.31 | 0 | 9 |  |
| Anxiety Severity | 18.83 | 18.28 | 0 | 100 | 2288 |
| Depression Severity | 29.26 | 19.01 | 0 | 100 | 2288 |
| Waist to Hip Ratio | 0.91 | 0.10 | 0.40 | 1.29 | 1911 |
| BMI | 30.64 | 8.30 | 7.9 | 100.3 | 1967 |
| Obese | 0.48 | 0.50 | 0 | 1 | 1967 |
| Height | 169.44 | 11.28 | 105 | 199.5 | 1983 |

Note. Survey weights were applied to adjust the descriptive statistics. The sample size is 2,345 when the sample size column is missing,

**Table S2.** The distribution of network size and the proportion of hasslers.

| **Network**<br>**Size** | **Freq** | **Percent** | **%**<br>**Hasslers** |
| --- | --- | --- | --- |
| 0 | 35 | 1.3 |  |
| 1 | 97 | 3.6 | 7.2 |
| 2 | 232 | 8.6 | 6.9 |
| 3 | 415 | 15.5 | 6.8 |
| 4 | 461 | 17.2 | 9.4 |
| 5 | 404 | 15 | 8.3 |
| 6 | 343 | 12.8 | 7.8 |
| 7 | 247 | 9.2 | 8.8 |
| 8 | 139 | 5.2 | 9.5 |
| 9 | 80 | 3 | 7.4 |
| 10+ | 232 | 8.6 | 8.4 |
| Total | 2685 | 100 |  |
| mean | 5.07 |  | 8.1 |
| sd | 2.46 |  | 15.9 |

**Table S3.**
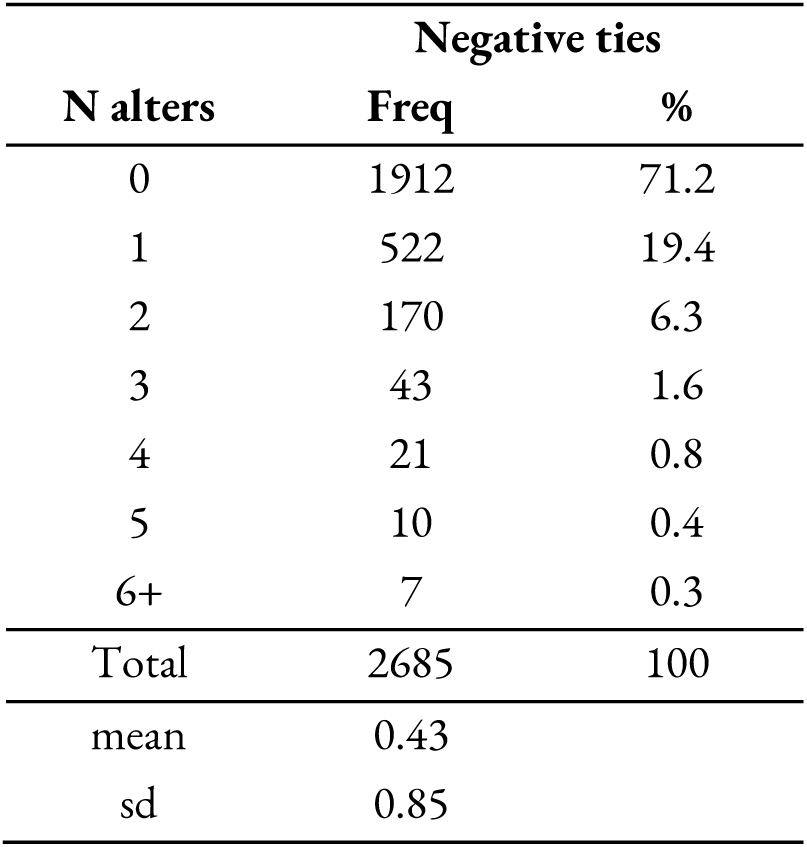
The distribution of the size of negative ties.

| **N alters** | **Negative ties** |  |
| --- | --- | --- |
|  | **Freq** | **%** |
| 0 | 1912 | 71.2 |
| 1 | 522 | 19.4 |
| 2 | 170 | 6.3 |
| 3 | 43 | 1.6 |
| 4 | 21 | 0.8 |
| 5 | 10 | 0.4 |
| 6+ | 7 | 0.3 |
| Total | 2685 | 100 |
| mean | 0.43 |  |
| sd | 0.85 |  |

**Table S4.** Regression results from the zero-inflated Poisson regression model for the presence and the number of hasslers in ego networks.

| Outcome | Model Coefficients |  | Average Marginal Effects |  |
| --- | --- | --- | --- | --- |
|  | Zero Hasslers | Count of Hasslers | Zero Hasslers | Count of Hasslers |
| Age (SD) | 0.400<br>(0.242) | 0.0667<br>(0.107) | 0.0243+<br>(0.0142) | -0.0272<br>(0.0305) |
| Network size (SD) | -0.234<br>(0.226) | 0.447**<br>(0.0508) | -0.0942**<br>(0.0134) | 0.219**<br>(0.0222) |
| Self-reported general health (SD) | -0.175<br>(0.230) | -0.271*<br>(0.118) | 0.0292*<br>(0.0114) | -0.0893**<br>(0.0280) |
| Lifetime Multimorbidity Index (SD) | -0.483<br>(0.366) | -0.148<br>(0.106) | -0.0182<br>(0.0216) | 0.00462<br>(0.0251) |
| ACEs score (SD) | -0.711**<br>(0.182) | 0.0715<br>(0.0604) | -0.0743**<br>(0.0112) | 0.128**<br>(0.0195) |
| Health Insurance | -1.132<br>(0.720) | -0.354<br>(0.280) | -0.0415<br>(0.0297) | 0.00772<br>(0.0735) |
| Survey during COVID-19 | -0.181<br>(0.821) | -0.300<br>(0.286) | 0.0333<br>(0.0360) | -0.100*<br>(0.0469) |
| How much do others depend on you | -0.677+<br>(0.376) | 0.0331<br>(0.127) | -0.0651*<br>(0.0283) | 0.107+<br>(0.0534) |
| How important are you to others | 0.955<br>(0.612) | 0.0134<br>(0.190) | 0.0819**<br>(0.0254) | -0.126**<br>(0.0435) |
| Education (ref = College or higher) |  |  |  |  |
| HS or less than HS | 0.358<br>(0.569) | 0.230<br>(0.245) | -0.00969<br>(0.0343) | 0.0510<br>(0.0706) |
| Some college | 0.769<br>(0.525) | 0.253<br>(0.231) | 0.0254<br>(0.0347) | -0.00176<br>(0.0695) |
| Race (ref = White) |  |  |  |  |
| Black | 0.357<br>(0.843) | 0.192<br>(0.315) | 0.00269<br>(0.0573) | 0.0279<br>(0.105) |
| Other | -0.259<br>(0.904) | 0.117<br>(0.190) | -0.0435<br>(0.0845) | 0.0891<br>(0.151) |
| Female (vs male) | -0.779<br>(0.603) | -0.0559<br>(0.201) | -0.0610*<br>(0.0266) | 0.0863*<br>(0.0424) |
| Marital Status (ref = Never-married) |  |  |  |  |
| Married or living with a partner | 0.0891<br>(0.673) | 0.226<br>(0.283) | -0.0286<br>(0.0336) | 0.0805<br>(0.0700) |
| Widowed/Divorced/Separated | -0.290 | -0.0286 | -0.0184 | 0.0235 |
|  | (0.746) | (0.314) | (0.0309) | (0.0591) |
| Occupation (ref = Unemployed) |  |  |  |  |
| Professional/Managerial | 0.288<br>(1.657) | -0.211<br>(0.478) | 0.0690<br>(0.132) | -0.183<br>(0.197) |
| Healthcare | 0.453<br>(1.378) | -0.570+<br>(0.316) | 0.140<br>(0.113) | -0.357*<br>(0.161) |
| Education & Social Services | 0.462<br>(1.476) | -0.812*<br>(0.387) | 0.174<br>(0.118) | -0.430*<br>(0.176) |
| Service | 0.700<br>(1.513) | -0.208<br>(0.379) | 0.112<br>(0.130) | -0.255<br>(0.197) |
| Sales & Office | 0.389<br>(1.364) | -0.421<br>(0.385) | 0.112<br>(0.115) | -0.294<br>(0.185) |
| Production & Transportation | -22.02**<br>(3.752) | -1.124**<br>(0.409) | 0.0622<br>(0.131) | -0.310<br>(0.207) |
| Other | -0.183<br>(1.500) | -0.519<br>(0.451) | 0.0779<br>(0.118) | -0.257<br>(0.196) |
| Not in labor force | -0.661<br>(1.162) | -0.747**<br>(0.271) | 0.0849<br>(0.101) | -0.300+<br>(0.163) |
| Missing Industry | 0.314<br>(1.415) | -0.394<br>(0.367) | 0.101<br>(0.113) | -0.272<br>(0.166) |
| Smoking status (ref = never smoker) |  |  |  |  |
| ever smoker | 0.255<br>(0.519) | -0.0139<br>(0.177) | 0.0241<br>(0.0329) | -0.0384<br>(0.0542) |
| current smoker | 1.057<br>(0.779) | 0.318<br>(0.388) | 0.0534<br>(0.0505) | -0.0369<br>(0.118) |
| daily smoker | -0.361<br>(0.648) | 0.221<br>(0.306) | -0.0728*<br>(0.0339) | 0.157+<br>(0.0902) |
| Constant | 0.431<br>(1.340) | 0.0755<br>(0.393) |  |  |
| Observations | 2596 |  | 2596 | 2596 |

Note. We estimate a Zero-inflated Poisson regression model using Stata's `svy: zip` command to account for the complex survey design. We report both regression coefficients and average marginal effects for the probability of zero hasslers ( $\Pr(0)$ ) and for the expected count of hasslers. Standard errors are shown in parentheses (+ $p < 0.1$ , \* $p < 0.05$ , \*\* $p < 0.01$ ).

**Table S5.** OLS regression models predicting two epigenetic aging clocks.

| Outcome | PACE |  |  |  | GrimAge2 |  |
| --- | --- | --- | --- | --- | --- | --- |
|  | Model1 | Model2 | Model3 | Model4 | Model5 | Model6 |
| Batch Number (ref = 8615) |  |  |  |  |  |  |
| 8732 | -0.0283** | -0.0292** | -0.0288** | -0.545 | -0.590 | -0.554 |
|  | (0.00804) | (0.00806) | (0.00783) | (0.478) | (0.472) | (0.481) |
| 9054 | -0.0301** | -0.0297** | -0.0282** | 0.124 | 0.142 | 0.162 |
|  | (0.0101) | (0.00997) | (0.00976) | (0.318) | (0.329) | (0.316) |
| 9109 | -0.00756 | -0.00781 | -0.00729 | 0.0853 | 0.0759 | 0.0873 |
|  | (0.0101) | (0.0103) | (0.00978) | (0.448) | (0.444) | (0.439) |
| 9213 | -0.0539** | -0.0544** | -0.0526** | -0.397 | -0.425 | -0.371 |
|  | (0.0122) | (0.0122) | (0.0121) | (0.481) | (0.499) | (0.486) |
| % Leukocyte cells | 0.0407 | 0.0397 | 0.0398 | 0.375 | 0.329 | 0.378 |
|  | (0.0452) | (0.0452) | (0.0452) | (0.803) | (0.830) | (0.769) |
| Age | -0.0474+ | -0.0485+ | -0.0463+ | -0.624 | -0.684 | -0.610 |
|  | (0.0244) | (0.0243) | (0.0244) | (0.997) | (0.997) | (0.998) |
| Race (ref = White) | -0.676** | -0.680** | -0.681** | -26.46** | -26.65** | -26.55** |
| Black | (0.0347) | (0.0344) | (0.0353) | (0.984) | (0.982) | (0.987) |
|  | 0.000790* | 0.000783* | 0.000827* |  |  |  |
|  | * | * | * | -0.0476** | -0.0482** | -0.0468** |
|  | (0.000276 | (0.000279 | (0.000276 |  |  |  |
| Other | ) | ) | ) | (0.00762) | (0.00777) | (0.00770) |
| FEMALE | 0.0807** | 0.0819** | 0.0821** | 2.273** | 2.337** | 2.294** |
|  | (0.0125) | (0.0125) | (0.0132) | (0.502) | (0.523) | (0.525) |
| Education (ref = HS or less than HS) | 0.0532** | 0.0558** | 0.0543** | 1.454** | 1.591** | 1.433** |
| Some college | (0.00948) | (0.0105) | (0.0100) | (0.487) | (0.564) | (0.476) |
|  | 0.0303** | 0.0303** | 0.0301** | -0.266 | -0.259 | -0.269 |
| College or higher | (0.00576) | (0.00574) | (0.00564) | (0.227) | (0.227) | (0.231) |
| Marital Status (ref = Never-married) | -0.0244* | -0.0250* | -0.0243* | -0.969* | -1.006* | -0.957* |
| Married or living with a partner | (0.0116) | (0.0116) | (0.0119) | (0.424) | (0.421) | (0.429) |
|  | -0.0599** | -0.0615** | -0.0599** | -2.824** | -2.913** | -2.816** |
| Widowed/Divorced/Separated | (0.00943) | (0.00936) | (0.00959) | (0.462) | (0.456) | (0.468) |
| Number of hasslers | -0.00654 | -0.00666 | -0.00539 | 0.583+ | 0.585+ | 0.597+ |
|  | (0.0120) | (0.0123) | (0.0121) | (0.318) | (0.328) | (0.309) |
| Presence of any hassler | 0.0185 | 0.0179 | 0.0175 | 1.806** | 1.782** | 1.782** |
|  | (0.0125) | (0.0126) | (0.0122) | (0.423) | (0.429) | (0.421) |
| N Hasslers (ref = 0) | 0.0152** |  |  | 0.782** |  |  |
| 1 | (0.00382) |  |  | (0.172) |  |  |
|  |  | 0.0259** |  |  | 1.252** |  |
| 2 |  | (0.00732) |  |  | (0.290) |  |
| 3 |  |  | 0.0110 |  |  | 0.707* |
|  |  |  | (0.00782) |  |  | (0.282) |
| 4 |  |  | 0.0681** |  |  | 2.339** |
|  |  |  | (0.0157) |  |  | (0.340) |
| 5+ |  |  | 0.0444 |  |  | 1.721 |
|  |  |  | (0.0362) |  |  | (1.159) |
| Constant |  |  | 0.0961+ |  |  | 4.494** |
|  |  |  | (0.0523) |  |  | (1.076) |
| Observations |  |  | -0.0126 |  |  | 2.615 |

Note. All models control for network size dummies. Standard errors are in parentheses (+ p < 0.1, \* p < 0.05, \*\* p < 0.01).

**Table S6.** Results from the alternative specification of negative ties.

|  | PACE |  |  | AgeAccelGrim2 |  |  |
| --- | --- | --- | --- | --- | --- | --- |
|  | Model1 | Model2 | Model3 | Model4 | Model5 | Model6 |
| % Negative Ties | 0.0793**<br>(0.0208) |  |  | 4.059**<br>(0.985) |  |  |
| % Hasslers (Ref = 0%) |  |  |  |  |  |  |
| % Hasslers < 25% |  | 0.0141<br>(0.00993) |  |  | 1.031**<br>(0.265) |  |
| % Hasslers < 50% |  | 0.0368**<br>(0.0130) |  |  | 1.101**<br>(0.402) |  |
| % Hasslers < 100% |  | 0.0112<br>(0.0213) |  |  | 1.376<br>(0.845) |  |
| % Hasslers = 100% |  | 0.0846*<br>(0.0316) |  |  | 5.002**<br>(1.468) |  |
| Avg Hassling Frequency |  |  | 0.0148*<br>(0.00614) |  |  | 0.695**<br>(0.233) |
| Observations | 2345 | 2345 | 2345 | 2345 | 2345 | 2345 |
| Controls |  |  |  |  |  |  |
| Individual controls | V | V | V | V | V | V |
| Network size dummies | V | V | V | V | V | V |

**Table S7.** OLS regression models predicting self-reported health outcomes.

| Outcome | General Health | Mental Health | Physical Health | Anxiety Severity | Depression Severity |
| --- | --- | --- | --- | --- | --- |
| Models | Model1 | Model2 | Model3 | Model4 | Model5 |
| Bath number (ref = 8615) |  |  |  |  |  |
| 8732 | -0.0346<br>(0.0776) | -0.0219<br>(0.0979) | -0.0368<br>(0.0984) | -0.796<br>(1.697) | 0.254<br>(1.700) |
| 9054 | -0.0238<br>(0.0894) | 0.0153<br>(0.0960) | -0.0262<br>(0.102) | -0.314<br>(1.768) | 0.260<br>(1.809) |
| 9109 | 0.128<br>(0.0810) | 0.147<br>(0.106) | 0.190*<br>(0.0905) | 0.738<br>(1.823) | 1.535<br>(1.886) |
| 9213 | -0.00323<br>(0.107) | -0.00401<br>(0.0892) | -0.0409<br>(0.118) | 4.789*<br>(2.031) | 2.848<br>(1.791) |
| 11277 | -0.256<br>(0.261) | -0.127<br>(0.246) | -0.116<br>(0.290) | -7.323**<br>(2.612) | -10.04*<br>(4.355) |
| 13762 | -0.0991<br>(0.133) | -0.0656<br>(0.127) | -0.00820<br>(0.136) | -7.582**<br>(1.709) | -5.642*<br>(2.690) |
| % Leukocyte cells | -0.479**<br>(0.134) | -0.377*<br>(0.159) | -0.496**<br>(0.130) | -7.673**<br>(2.566) | -7.848**<br>(2.790) |
| Age | -0.000254<br>(0.00163) | -0.00978**<br>(0.00176) | 0.00263<br>(0.00167) | -0.208**<br>(0.0336) | -0.169**<br>(0.0348) |
| Race (ref = White) |  |  |  |  |  |
| Black | 0.0899<br>(0.0776) | -0.140*<br>(0.0612) | 0.000554<br>(0.0685) | -2.853*<br>(1.220) | -4.983**<br>(1.326) |
| Other | 0.127<br>(0.117) | -0.0272<br>(0.116) | 0.170<br>(0.159) | 0.770<br>(1.853) | -2.577<br>(1.960) |
| FEMALE | 0.125*<br>(0.0561) | 0.251**<br>(0.0486) | 0.223**<br>(0.0449) | 3.583**<br>(0.955) | 3.553**<br>(0.985) |
| Education (ref = HS or less than HS) |  |  |  |  |  |
| Some college | -0.228**<br>(0.0649) | -0.258**<br>(0.0795) | -0.260**<br>(0.0517) | -1.053<br>(1.596) | 0.0765<br>(1.232) |
| College or higher | -0.405**<br>(0.0749) | -0.393**<br>(0.0801) | -0.475**<br>(0.0652) | -1.995<br>(1.755) | -1.574<br>(1.300) |
| Marital Status (ref = Never-married) |  |  |  |  |  |
| Married or living with a partner | -0.149<br>(0.0894) | -0.0799<br>(0.0940) | -0.194*<br>(0.0880) | -2.273<br>(1.727) | -3.537*<br>(1.548) |
| Widowed/Divorced/Separated | 0.0274<br>(0.106) | 0.144<br>(0.118) | -0.0570<br>(0.103) | 1.580<br>(2.113) | 1.866<br>(1.929) |
| Constant | 3.016**<br>(0.363) | 3.392**<br>(0.314) | 2.808**<br>(0.408) | 37.52**<br>(8.035) | 45.43**<br>(9.196) |
| Observations | 2343 | 2342 | 2343 | 2288 | 2288 |

Note. All models control for network size dummies. Standard errors are in parentheses (+ $p < 0.1$ , \* $p < 0.05$ , \*\* $p < 0.01$ ).

**Table S8.** OLS regression models predicting objective health outcomes.

| Outcome | Epigenetic<br>inflammation<br>score | Multi-<br>morbidity | Waist to<br>Hip Ratio | BMI | Height |
| --- | --- | --- | --- | --- | --- |
| Models | Model6 | Model7 | Model8 | Model9 | Model10 |
| Bath number (ref = 8615) |  |  |  |  |  |
| 8732 | -0.00322**<br>(0.000763) | 0.00664<br>(0.124) | -0.00374<br>(0.0106) | -1.030<br>(0.861) | -0.928<br>(1.391) |
| 9054 | -0.00344**<br>(0.000746) | -0.00868<br>(0.0926) | -0.00799<br>(0.0118) | -1.065<br>(0.958) | 0.739<br>(1.149) |
| 9109 | -0.00207*<br>(0.000820) | 0.108<br>(0.122) | -0.00443<br>(0.00875) | -1.056<br>(0.751) | 0.354<br>(1.520) |
| 9213 | -0.00380**<br>(0.000964) | -0.0526<br>(0.115) | -0.00750<br>(0.0107) | -2.461*<br>(0.943) | 0.0589<br>(1.154) |
| 11277 | 0.00160<br>(0.00240) | -0.249<br>(0.289) | 0.0216<br>(0.0168) | 0.949<br>(2.372) | -0.504<br>(1.300) |
| 13762 | -0.00474*<br>(0.00180) | 0.152<br>(0.183) | -0.00751<br>(0.0136) | 0.715<br>(1.650) | 1.235<br>(2.046) |
| % Leukocyte cells | -0.146**<br>(0.00182) | -0.529+<br>(0.299) | 0.00865<br>(0.0107) | 2.019<br>(1.210) | -3.492**<br>(1.262) |
| Age | 0.000144**<br>(0.0000177) | 0.0270**<br>(0.00208) | 0.000880**<br>(0.000286) | 0.00139<br>(0.0162) | -0.102**<br>(0.0133) |
| Race (ref = White) |  |  |  |  |  |
| Black | 0.00352**<br>(0.00128) | 0.0360<br>(0.111) | 0.00868<br>(0.00907) | 1.399+<br>(0.726) | -1.328<br>(0.802) |
| Other | 0.00261*<br>(0.00116) | -0.0221<br>(0.116) | 0.00891<br>(0.0133) | 1.093<br>(1.096) | -2.429**<br>(0.740) |
| FEMALE | 0.00243**<br>(0.000446) | 0.118+<br>(0.0700) | -0.0784**<br>(0.00489) | 0.663<br>(0.444) | -14.52**<br>(0.577) |
| Education (ref = HS or less than HS) |  |  |  |  |  |
| Some college | -0.00152+<br>(0.000799) | -0.189+<br>(0.105) | -0.00905<br>(0.00839) | -0.0393<br>(0.824) | 0.566<br>(0.373) |
| College or higher | -0.00488**<br>(0.000955) | -0.199+<br>(0.106) | -0.0168**<br>(0.00582) | -1.328*<br>(0.637) | 1.682*<br>(0.730) |
| Marital Status (ref = Never-married) |  |  |  |  |  |
| Married or living with a partner | -0.000434<br>(0.000866) | -0.0287<br>(0.107) | 0.0294*<br>(0.0109) | 0.941<br>(0.899) | -0.534<br>(0.845) |
| Widowed/Divorced/Separated | 0.00283**<br>(0.000898) | -0.0317<br>(0.118) | 0.0287*<br>(0.0122) | 1.171<br>(1.221) | -0.957<br>(0.972) |
| Constant | 0.156**<br>(0.00326) | 0.0444<br>(0.347) | 0.912**<br>(0.0290) | 25.79**<br>(2.582) | 186.0**<br>(3.909) |
| Observations | 2345 | 2158 | 1911 | 1967 | 1983 |

Note. All models control for network size dummies. Standard errors are in parentheses (+ $p < 0.1$ , \* $p < 0.05$ , \*\* $p < 0.01$ ).

**Table S9.**
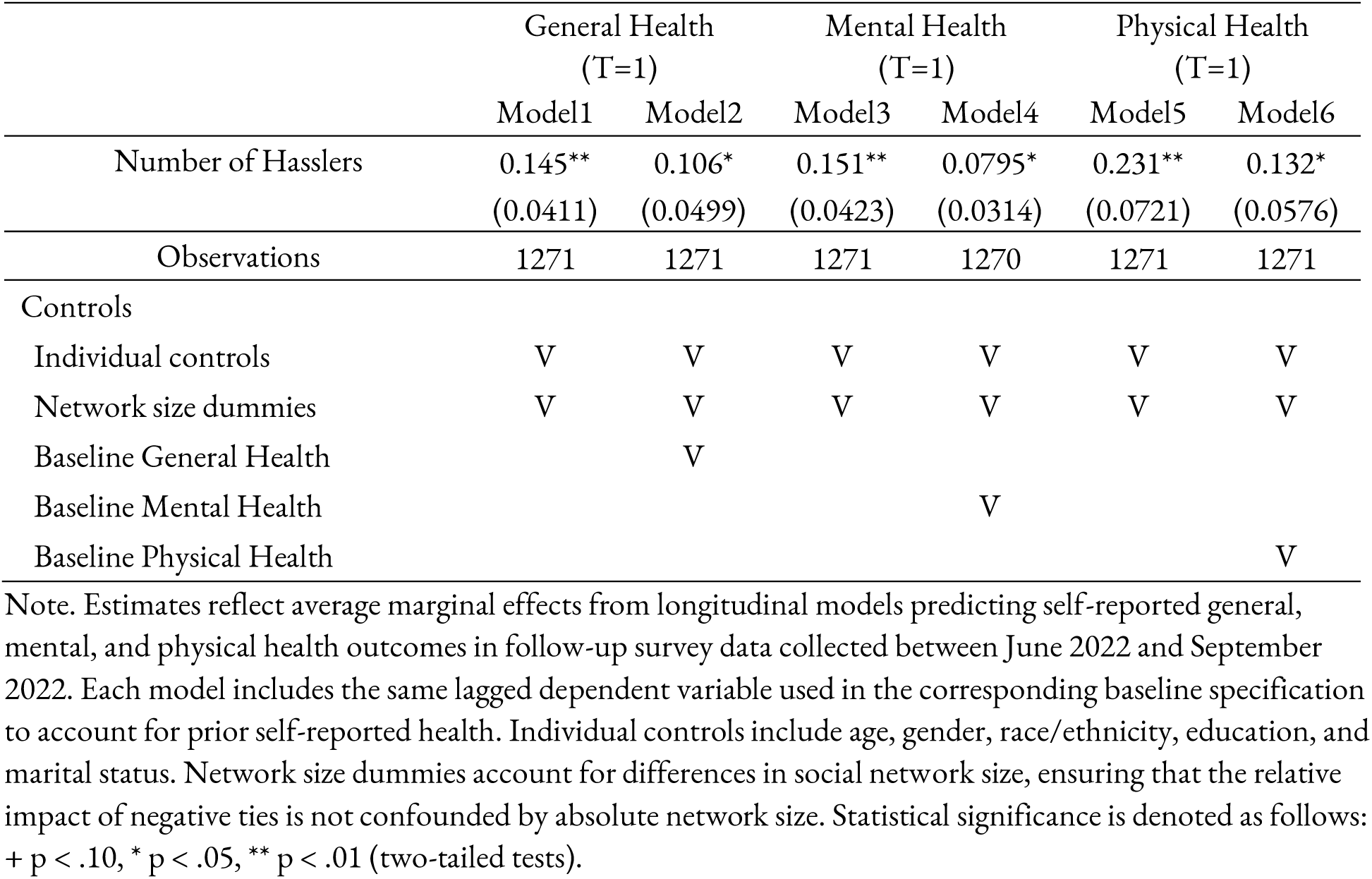
Longitudinal Analyses of the Association Between Negative Social Ties and Self-reported Health outcomes.

|  | General Health<br>(T=1) |  | Mental Health<br>(T=1) |  | Physical Health<br>(T=1) |  |
| --- | --- | --- | --- | --- | --- | --- |
|  | Model1 | Model2 | Model3 | Model4 | Model5 | Model6 |
| Number of Hasslers | 0.145**<br>(0.0411) | 0.106*<br>(0.0499) | 0.151**<br>(0.0423) | 0.0795*<br>(0.0314) | 0.231**<br>(0.0721) | 0.132*<br>(0.0576) |
| Observations | 1271 | 1271 | 1271 | 1270 | 1271 | 1271 |
| Controls |  |  |  |  |  |  |
| Individual controls | V | V | V | V | V | V |
| Network size dummies | V | V | V | V | V | V |
| Baseline General Health |  | V |  |  |  |  |
| Baseline Mental Health |  |  |  | V |  |  |
| Baseline Physical Health |  |  |  |  |  | V |

Note. Estimates reflect average marginal effects from longitudinal models predicting self-reported general, mental, and physical health outcomes in follow-up survey data collected between June 2022 and September 2022. Each model includes the same lagged dependent variable used in the corresponding baseline specification to account for prior self-reported health. Individual controls include age, gender, race/ethnicity, education, and marital status. Network size dummies account for differences in social network size, ensuring that the relative impact of negative ties is not confounded by absolute network size. Statistical significance is denoted as follows: + $p < .10$ , \* $p < .05$ , \*\* $p < .01$ (two-tailed tests).

**Table S10.** Sensitivity of associations between the number of hasslers and epigenetic aging outcomes to unmeasured confounding from *konfound* analysis.

|  | PACE | AgeAccelGrim2 |
| --- | --- | --- |
| **RIR cases** | **1088 (46.4%)** | **1129 (48.2%)** |
| threshold effect size | 0.007 | 0.313 |
| **Conditional ITCV for the number of hasslers** | **0.036** | **0.039** |
| Threshold correlation (confounder, outcome) | 0.191 | 0.198 |
| Threshold correlation (confounder, predictor) | 0.191 | 0.198 |
| Conditional ITCV for other covariates |  |  |
| Batch 2 (vs Batch 1) | 0.001 | 0.000 |
| Batch 3 (vs Batch 1) | 0.001 | 0.000 |
| Batch 4 (vs Batch 1) | 0.000 | 0.000 |
| Batch 5 (vs Batch 1) | 0.001 | 0.000 |
| Batch 6 (vs Batch 1) | 0.001 | 0.001 |
| Batch 7 (vs Batch 1) | 0.002 | 0.001 |
| Leukocytes | 0.034 | 0.037 |
| Age | -0.003 | 0.030 |
| Race : Black (Vs White) | 0.001 | 0.001 |
| Race : Other (Vs White) | 0.002 | 0.001 |
| Female (vs Male) | 0.006 | -0.007 |
| Educ: Some college (vs HS or less than HS) | 0.001 | 0.001 |
| Educ: College or higher (vs HS or less than HS) | 0.000 | 0.000 |
| Marital Status: Married or Cohabited (vs never-married) | 0.001 | 0.002 |
| Marital Status: Widowed/Divorced/Separated (vs never-married) | 0.003 | 0.006 |

Note. Values represent robustness statistics from *konfound* analyses evaluating the extent of unmeasured confounding required to eliminate the observed associations between the number of hasslers and two epigenetic aging measures (PACE and AgeAccelGrim2). RIR (Robustness of Inference to Replacement) indicates the percentage and number of cases that would need to be replaced with counterfactual cases showing null effects to render the association nonsignificant at $p < 0.05$ . ITCV (Impact Threshold for a Confounding Variable) reflects the minimum product of correlations that an omitted confounder would need with both the predictor and the outcome to explain away the effect; corresponding threshold correlations are shown. Conditional ITCV values for all measured covariates illustrate that none approach the threshold required to overturn the focal association.

**Table S11.** Associations Between Hasslers and Biological Aging Across Age Groups.

| Outcomes | Pace |  |  |  |  |  |
| --- | --- | --- | --- | --- | --- | --- |
| Age Group | 18-30 | 31-40 | 41-50 | 51-60 | 61-70 | 71+ |
| N of Hasslers | **0.0368\*\***<br>**(0.00837)** | 0.00126<br>(0.00834) | 0.0156<br>(0.0164) | -0.000233<br>(0.00733) | 0.00328<br>(0.00859) | 0.00915<br>(0.0118) |
| Observations | 375 | 425 | 339 | 370 | 407 | 363 |
| Outcomes | AgeAccelGrim2 |  |  |  |  |  |
| Age Group | 18-30 | 31-40 | 41-50 | 51-60 | 61-70 | 71+ |
| N of Hasslers | 0.953**<br>(0.288) | 0.322<br>(0.313) | 0.553+<br>(0.303) | 1.166**<br>(0.226) | 0.0525<br>(0.301) | 0.805+<br>(0.420) |
| Observations | 375 | 425 | 339 | 370 | 407 | 363 |

Note. Coefficients represent the average marginal effects of negative social ties on biological aging outcomes, with standard errors in parentheses. Individual controls include age, gender, race/ethnicity, education, and marital status. Network size dummies account for differences in social network size, ensuring that the relative impact of negative ties is not confounded by absolute network size. Statistical significance is denoted as follows: + $p < .10$ , \* $p < .05$ , \*\* $p < .01$ (two-tailed tests).

**Figure S1.**
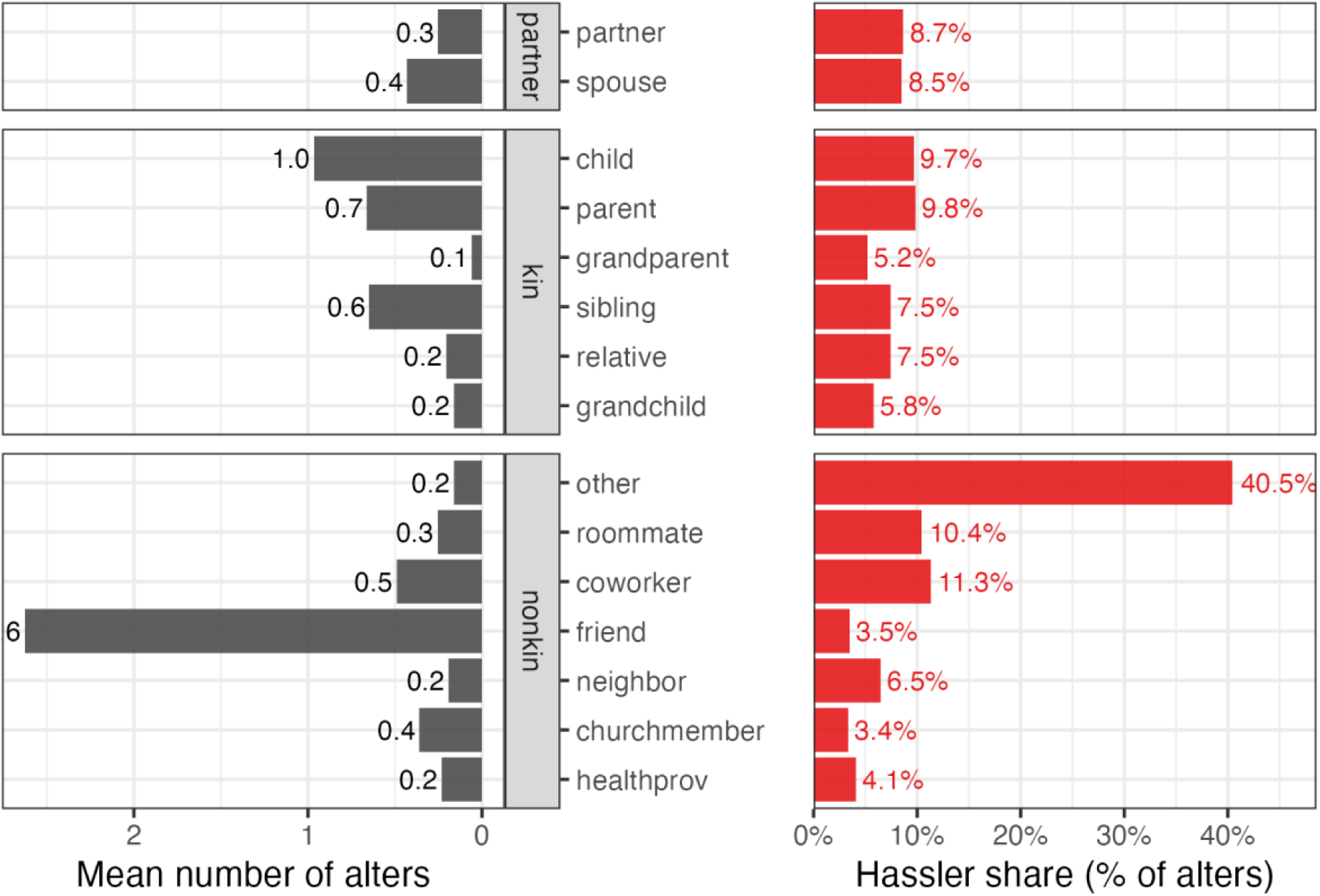
The mean number of alters by relationship type and the proportion of hasslers in each relationship category. Note. Mean number of alters (left panels) and proportion of alters who are reported as “hasslers” (right panels) across partner, kin, and non-kin relationship categories. Bars show the number of each alter type (e.g., spouse, parent, coworker), and the percentage (i.e., the share of alters within each category who are identified as hasslers). All values reflect the average values across all respondents.

